# Evaluating Genomic Surveillance Methods for *Shigella sonnei* in a High-Income Setting

**DOI:** 10.64898/2026.05.08.26352707

**Authors:** Kuangyi Charles Wei, Charlotte E. Chong, Gherard Batisti Biffignandi, Lewis C. E. Mason, Ryan Morrison, Claire Jenkins, Kate S Baker

## Abstract

*Shigella sonnei* is a human-adapted enteric pathogen with a very low infectious dose and increasing antimicrobial resistance. In high-income settings, transmission is multimodal including sporadic cases/outbreaks associated with food and travel, as well as sustained transmission among sexual networks of men who have sex with men (MSM). Whole-genome sequencing (WGS) now underpins national shigellosis surveillance in the United Kingdom. Hence, consistent, communicable genotyping is essential for case linkage and trend detection across heterogeneous transmission modes.

Here, we evaluate the performance of WGS genotyping approaches for granulating outbreaks of *S. sonnei* shigellosis, particularly considering differential performance in dense sexual transmission where highly clonal MSM-associated sublineages pose distinct clustering challenges. Specifically, we compare performance of the current practice approach (10 SNP-distance clustering based on SNP address [t10]), allele-based methods (EnteroBase cgMLST/HierCC [HC5]), a pathogen-specific genotyping scheme (sonneityper), and two k-mer based approaches (PopPUNK and KPop), on a *bona fide* UK surveillance dataset (n = 3,639 isolates from between 2016 and 2022), and stratify analyses by demographics (i.e. presumptive MSM [pMSM] versus non-pMSM).

Comparison metrics indicate that t10 clustering method groups data more broadly than HC5, and k-mer-based methods may capture genetic variation independent from SNP or allele-based approaches. Clusters derived from k-mer-based methods offer similar resolution to HC5 and reflect different demographics, but had unconvincing utility for this pathogen. These findings suggest a transmission context-aware surveillance workflow for shigellosis in high income settings: anchor routine communication on a portable allele-based backbone and augment with more granular, complementary methods (e.g., k-mer-based micro-partitioning or phylogenetic analysis) in comparatively low genomic-density regions of population structure (e.g., pMSM transmission lineages) to stabilise clusters and reduce artefactual chaining.

## Background

*Shigella* spp. are a major bacterial cause of diarrhoeal disease worldwide, responsible for an estimated 270 million episodes in people older than 5 years, 63,700 deaths among children younger than five years old, and 212,000 deaths across all ages annually (Khalil et al., 2018; WHO, 2022). Transmission occurs primarily via the faecal-oral route, and infections (shigellosis) are highly contagious, with an infectious dose of only 10-100 organisms (DuPont et al., 1989; Niyogi, 2005). Historically, shigellosis has been associated with contaminated food and water and person-to-person transmission, especially where hygiene and sanitation conditions are suboptimal, causing endemic disease in young children in low- and middle-income countries (LMICs). In high-income countries, shigellosis has been associated with travel to endemic regions, however, more recently *Shigella* spp. (primarily *S. flexneri* and *S. sonnei*) have emerged as sexually transmissible pathogens among men who have sex with men (MSM), with recurrent outbreaks reported since the early 2000s (Mason et al., 2024; Morgan et al., 2006; Siddiq et al., 2023; UKHSA, 2024). These two distinct transmission pathways, sporadic food- and travel-associated cases, and transmission through MSM sexual networks, pose different epidemiological (and hence evolutionary) scenarios that require tailored genomic surveillance strategies.

MSM-associated shigellosis outbreaks have coincided with the rise of antimicrobial resistance (AMR). Multi-drug resistant *Shigella* strains are now common, and in late 2021 an extensively drug-resistant (XDR) *S. sonnei* clade (CipR.MSM5, also known as Lineage 3, Clade 5) emerged among MSM in England, carrying plasmid-borne extended-spectrum β-lactamase (*bla*CTX-M-27) (Charles et al., 2022; Mason et al., 2023). This event, and the subsequent emergence of other XDR *Shigella* strains, underscore the urgent need for effective monitoring of AMR *Shigella* spp. in sexual transmission networks, where strains can spread rapidly (Gaudreau et al., 2022; Hawkey et al., 2021; Ingle et al., 2019; Lefèvre et al., 2023; Mason et al., 2023; Scott et al., 2025; Tansarli et al., 2023; Thorley et al., 2023). Public health interventions depend on timely detection of such outbreaks, yet even with whole-genome sequencing (WGS), distinguishing emerging clusters within intense transmission scenarios is challenging, owing to the close genetic distances among case isolates.

As such, highly discriminative WGS technology has transformed *Shigella* spp. surveillance, enabling high-resolution subtyping and outbreak detection (Armstrong et al., 2019; Dallman et al., 2015; den Bakker et al., 2014). Around the world, multiple genotyping approaches are already in use (Charles et al., 2024; ECDC, 2019; Nadon et al., 2017; WHO, 2023a, 2023b, 2018), which can be broadly classified into SNP- and allele- (most commonly cgMLST; core genome multilocus sequence typing) distance based methods. Meanwhile, k-mer-based methods are emerging as an alternative (Moore et al., 2024; Roberts et al., 2025; Rozday, 2025; Shi et al., 2025). Each approach has distinct strengths and limitations. SNP-based schemes offer fine-scale resolution (Baker et al., 2023; Dallman et al., 2018; Katz et al., 2017; Mixão et al., 2025) but can over-connect clusters through single-linkage chaining, as seen in the recent international XDR emergence among MSM where new XDR cases were obscured within a large pre-existing SNP-cluster (Mason et al., 2023). Allele-based approaches provide portable nomenclature and harmonisation across laboratories but may lack resolution for detecting emerging variants in clonal species like *S. sonnei* (Achtman et al., 2022; Jolley et al., 2018; Maiden et al., 2013; Mellmann et al., 2011; Zhou et al., 2021). Both traditional approaches omit the accessory genome (the large cloud of genetic diversity that is unstably present in a species [McInerney et al., 2017]). And while k-mer-based methods use all genomic information and are fast and scalable, they can be discordant with existing methods and may struggle to separate closely related isolates in low-diversity populations (Moore et al., 2024; Roberts et al., 2025; Rozday, 2025; Shi et al., 2025).

To address the relative efficacy of these approaches for effective surveillance of the multimodal transmission observed for *S. sonnei* in high income settings we systematically compared these genotyping approaches using *S. sonnei* isolates (n = 3,639) from England’s routine national surveillance programme from 2016 to 2022. We generated data for the isolates using an *S. sonnei* specific SNP-based Genotyping scheme implemented in Mykrobe (Hawkey et al., 2021), SNP-based clustering from an in-house UKHSA database (Dallman et al., 2018), cgMLST (Achtman et al., 2022), and two new k-mer-based methods, PopPUNK (Lees et al., 2019; Zhao et al., 2023) and KPop (Didelot and Ribeca, 2025). We analysed cross scheme concordance and used epidemiological metadata to assess which schemes best resolve transmission clusters in the two key contexts: sexual transmission and travel- and foodborne associated disease. In doing so, we demonstrate the strengths and limitations of each approach and propose guidelines for choosing appropriate genotyping strategies to support surveillance of the distinct public health scenarios.

## Results

### A high-quality genomic surveillance dataset of *S. sonnei*

We analysed 3,639 *Shigella sonnei* isolates sequenced between January 2016 and December 2022 as part of UKHSA’s routine genomic surveillance. After quality control (QC), 3,323 (91.3%) genomes were retained for downstream analyses. Exclusions were applied sequentially based on read-level, species-composition, and assembly-level QC, as well as the availability of essential genotyping outputs (**Methods**, **Figure 1a**).

**Figure 1:**
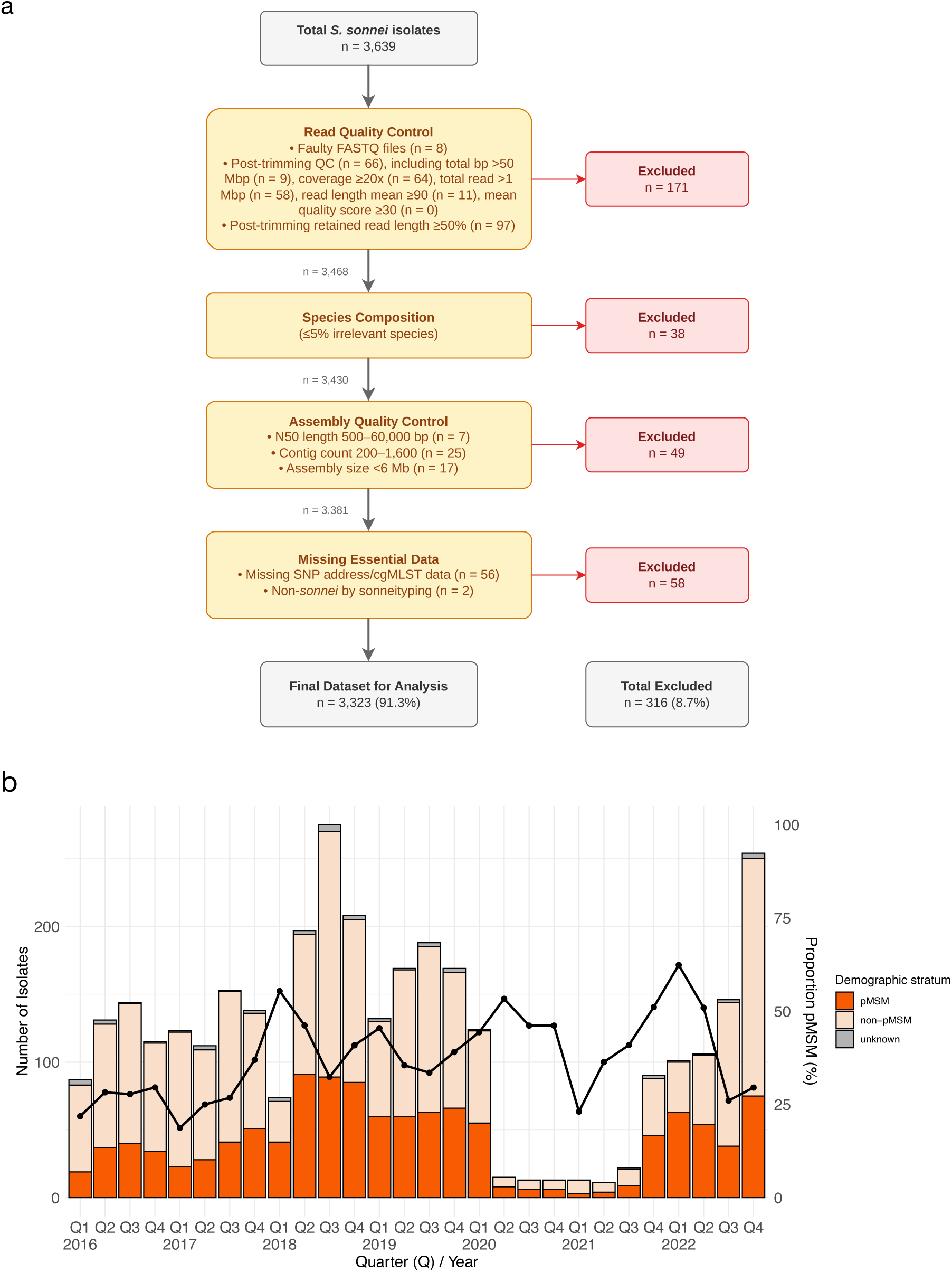
The dataset under study. **(a)** Quality control flowchart for the genomic data obtained from the *S. sonnei* routine surveillance. QC steps (yellow boxes) were applied sequentially according to arrows with exclusions removed (red arrows/boxes). Sample numbers (n) between boxes and final dataset represent remaining samples. **(b)** Breakdown of dataset over time and demographic groups shown as: isolate counts by quarter coloured according to the inlaid key (stacked bar chart, primary/left vertical axis) and proportion of isolates derived from pMSM over time (black line, secondary/right vertical axis).

Sampling was uneven over time, reflecting the underlying epidemiology (**Figure 1b**). Quarterly isolate counts ranged from 11 to 275, with notable peaks in late 2018 and winter 2022 coinciding with known increases in MSM-associated transmission of drug-resistant strains. A marked reduction in sampling occurred during 2020 and the first three quarters of 2021, reflecting disruptions to transmission, healthcare seeking and access, and routine surveillance during the COVID-19 pandemic. We stratified cases into presumptive MSM transmission (pMSM) and non-pMSM based on epidemiological metadata as previously, whereby adult males without recent history of travel to LMICs are defined as pMSM (see **Methods** [Mitchell et al., 2019, 2021; Marshall et al., 2026]). Of the high-quality genomes, 1,195 (36.0%) were classified as pMSM and 2,077 (62.5%) as non-pMSM. The proportion of isolates arising from pMSM increased over time (Cochran-Armitage trend, p < 0.001, from 21.8% in early 2016 to 29.5% in late 2022).

### *S. sonnei* grouping generation, including assessing k-mer based methods

As we aimed to compare a swathe of genomic surveillance approaches for shigellosis subtyping in multimodal transmission setting, we ran several well-established subtyping approaches for all 3,323 isolates. Specifically, the in-house SnapperDB (Dallman et al., 2018) which gave rise to 2,634 unique subtypes (called SNP addresses). SnapperDB works as a hierarchical single linkage clustering method at descending SNP distance thresholds. Specifically, the SNP address represents a sequential line of integers distinguishing clusters separated by a given number of SNPs, i.e. 250.100.50.25.10.5.0, with the routine practice for grouping *Shigella* for routine surveillance being used at t10 (i.e. 250.100.50.25.10 assigning 1,096 unique clusters) and t5 (i.e. 250.100.50.25.10.5 assigning 1,475 clusters). Next we ran a global genotyping framework (hereafter Genotyping; Hawkey et al., 2021) which returned 59 unique subtypes (called Genotypes), and finally we retrieved cgMLST subtypes from Enterobase (Achtman et al., 2022) defined using HierCC (Zhou et al., 2021) and employed the HC5 granularity (called HC5) indicating a distance of ≤ 5 allelic differences across the cgMLST loci (returning 1,153 unique HC5 clusters).

To explore the use of k-mer based methods, we implemented two k-mer tools, PopPUNK (Lees et al., 2019) and KPop (Didelot and Ribeca, 2025), on the dataset to produce clusters for comparison with the subtypes.

PopPUNK is a fast tool for bacterial genomic epidemiology that clusters genomes based on both core genome sequence divergence (π) and accessory genome content distance (a), using k-mer comparisons to estimate distances in lieu of computationally intensive genome alignment (Lees et al., 2019). Using PopPUNK on our dataset, we found the joint distribution of core distance (π) and accessory distance (a) was strongly concentrated at very small core distance (π = 0-1.5×10^−3^) with a low accessory distance (∼0-0.12), plus two high accessory distance islands (∼0.06-0.12;∼0.18-0.22) spanning a broad core distance range (**Supplementary Figure 1**). Clustering methods, including Hierarchical Density-Based Spatial Clustering of Applications with Noise (HDBSCAN) and Bayesian Gaussian Mixture Models (BGMM) on the core and accessory genome distance matrix did not yield clear group boundaries. Specifically, HDBSCAN produced hundreds of small, weakly connected components (components = 406, density = 0.0077, transitivity = 0.0089), whereas BGMM (K = 2-10) tended to remove most (43-98%, n = 1,435-3,247) samples and merge dense regions into a few large, highly connected clusters (e.g., when K = 5, components = 19, density = 0.2798, transitivity = 0.7269, see **Supplementary Table 1**). To assess performance for different modes of transmission, we then split our data demographically, which showed different low-distance kernels for the primary PopPUNK cluster for each group. Specifically, in pMSM, the highest-density contour was close to circular and in non-pMSM it was comparatively elongated over the axis of core distance (**Figure 2a-b**), possibly reflecting closer genetic relationships among sexually transmitting *Shigella*. Finally, PopPUNK lineage mode offered little improvement (**Supplementary Figure 2**). Overall, PopPUNK under-resolved structure in this highly clonal pathogen and did not provide stable, well-separated clusters.

**Figure 2:**
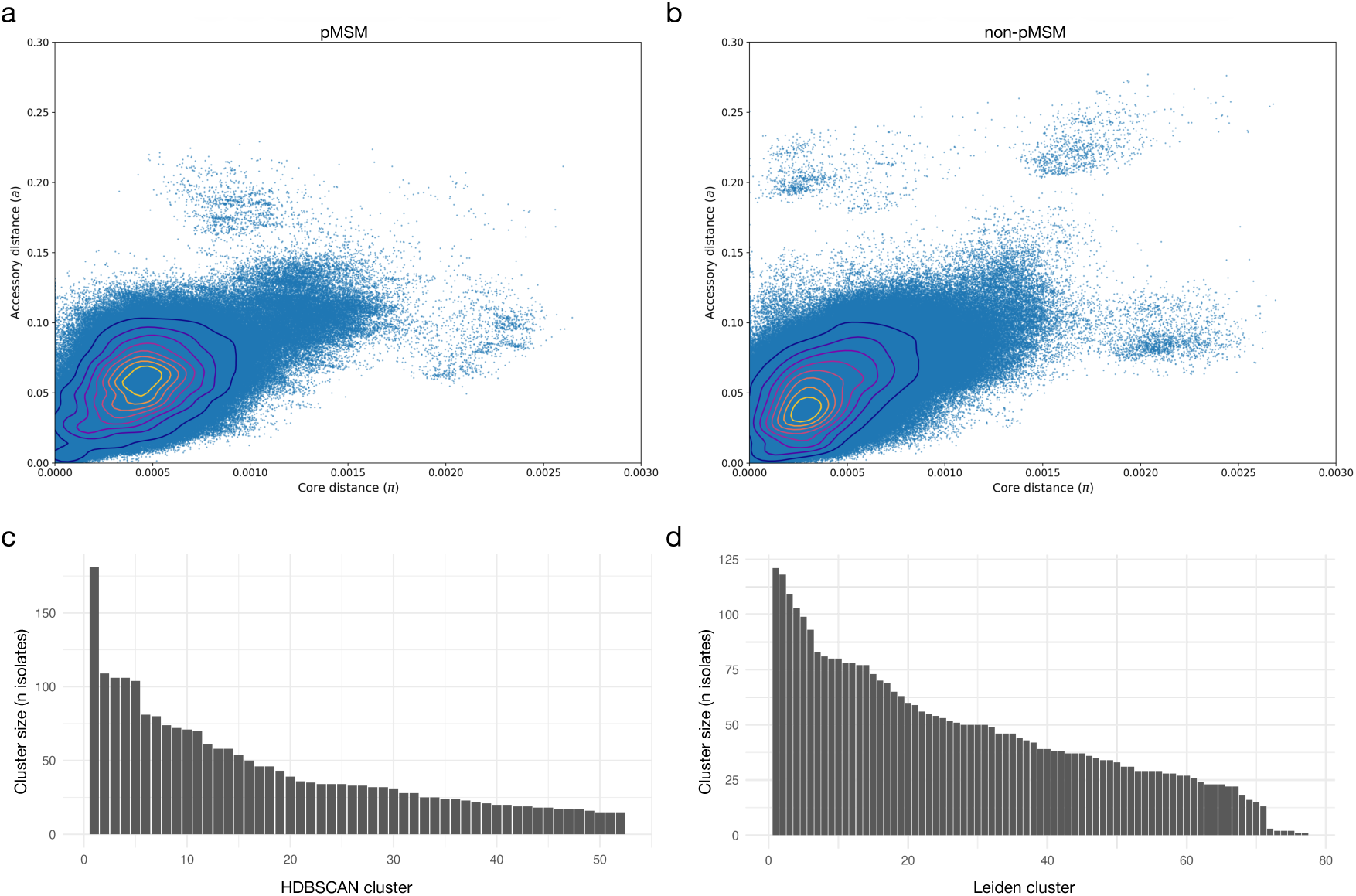
Clustering using the k-mer based methods PopPUNK and KPop. **(a-b)** PopPUNK pairwise distances for all *S. sonnei* samples stratified by pMSM **(a)** and non-pMSM **(b)** presented as a scatter of accessory distance (a) versus core distance (π). Points are overlaid with 2-D kernel-density contours highlighting the dense low-distance kernel. **(c-d)** Distribution of cluster size for the assigned non-singleton clusters from HDBSCAN **(c)** and Leiden **(d)** clustering using KPop distance matrix (cluster number is arbitrary from largest to smallest size).

An alternative k-mer based comparison method is KPop, a tool that represents each genome based on its complete k-mer composition. Instead of relying on sketches or predefined markers, KPop applies a correspondence analysis transformation to the k-mer profiles, producing compact numerical vectors that capture overall sequence composition (Didelot and Ribeca, 2025). These embeddings preserved the geometry of the original data, allowing for computation of meaningful distances between samples. A neighbour-joining tree generated from the distances showed little hierarchical structure with a star-like pattern rather than distinct clades and little correlation to other typing methods (i.e. Genotype, HC5, t10) (**Supplementary Figure 3**), so we similarly used clustering to facilitate scheme performance.

Applying HDBSCAN to the KPop distances revealed an expected trade-off between cluster fragmentation and conservatism. As the minimum cluster size (minPts) increased, the number of clusters fell (e.g., 705 at minPts = 2, 4 at minPts = 200), the average silhouette rose (0.47 to 0.64), and the fraction marked as noise increased (0.27 at minPts = 2, 0.52 at minPts = 200). Settings with minPts = 25-50 produced 17-31 clusters with mean silhouette 0.59-0.60 and noise fractions 0.33-0.36. We selected minPts = 15 because it jointly maximised silhouette mean and minimised noise fraction (0.35), yielding a moderate number of clusters (n = 52) and 1,149 isolates (34.6%) labelled as ‘noise’. In this scenario, the noise proportion may appropriately reflect unique subtypes and was comparable to the number of singletons produced by t10 (813, 24.5%) and HC5 (844, 25.4%) (**Supplementary Figure 4**). The corresponding distribution of cluster sizes is shown in **Figure 2c**. To examine the impact of clustering approach, we also employed Leiden clustering (which allows for more diverse cluster ‘shapes’) across a full parameter sweep on a k-nearest-neighbour graph constructed from the same KPop distance matrix. By nature, the Leiden approach assigned all samples to clusters (**Figure 2d**) so we optimised selection of the parameters (k and resolution) based on concordance with existing typing schema (t10 and HC5). Going forward, since HDBSCAN was not tuned against any external labels, we retain this as the primary k-mer-derived comparator in the main results (called KPop clusters, singletons-included) and the full Leiden parameter sweep and concordance analysis are available in the **Supplementary Material**.

### Cross-scheme agreement analysis

To assess the concordance between genotyping schemes on this dataset, we compared four schema, Genotyping, cgMLST/HierCC at the HC5 level, SNP-address at the t10 SNP single linkage cluster level, and KPop clusters (singletons-included), using a variety of uni- and bi-directional concordance measures (Adjusted Rand Index [ARI], Normalised Mutual Information [NMI], Adjusted Mutual Information [AMI], and directional Adjusted Wallace [AW]), globally and by demographic group (**Figure 3a-b**, **Supplementary Table 2**). Globally, the highest pairwise ARI was between HC5 and t10 (ARI = 0.644), followed by Genotype versus t10 (0.531) and Genotype versus HC5 (0.311); concordance of all methods with KPop clusters was poor (Genotype versus KPop at 0.109; HC5 versus KPop at 0.199; KPop versus t10 at 0.137). Directional AW revealed the nesting relationships: AW in the direction HC5 to t10 = 0.954, versus AW in the direction t10 to HC5 = 0.487, showed that HC5 groups were largely contained within t10 but that t10 often merges multiple HC5 communities, showing that HC5 is more discriminatory than t10 SNP-address.

**Figure 3:**
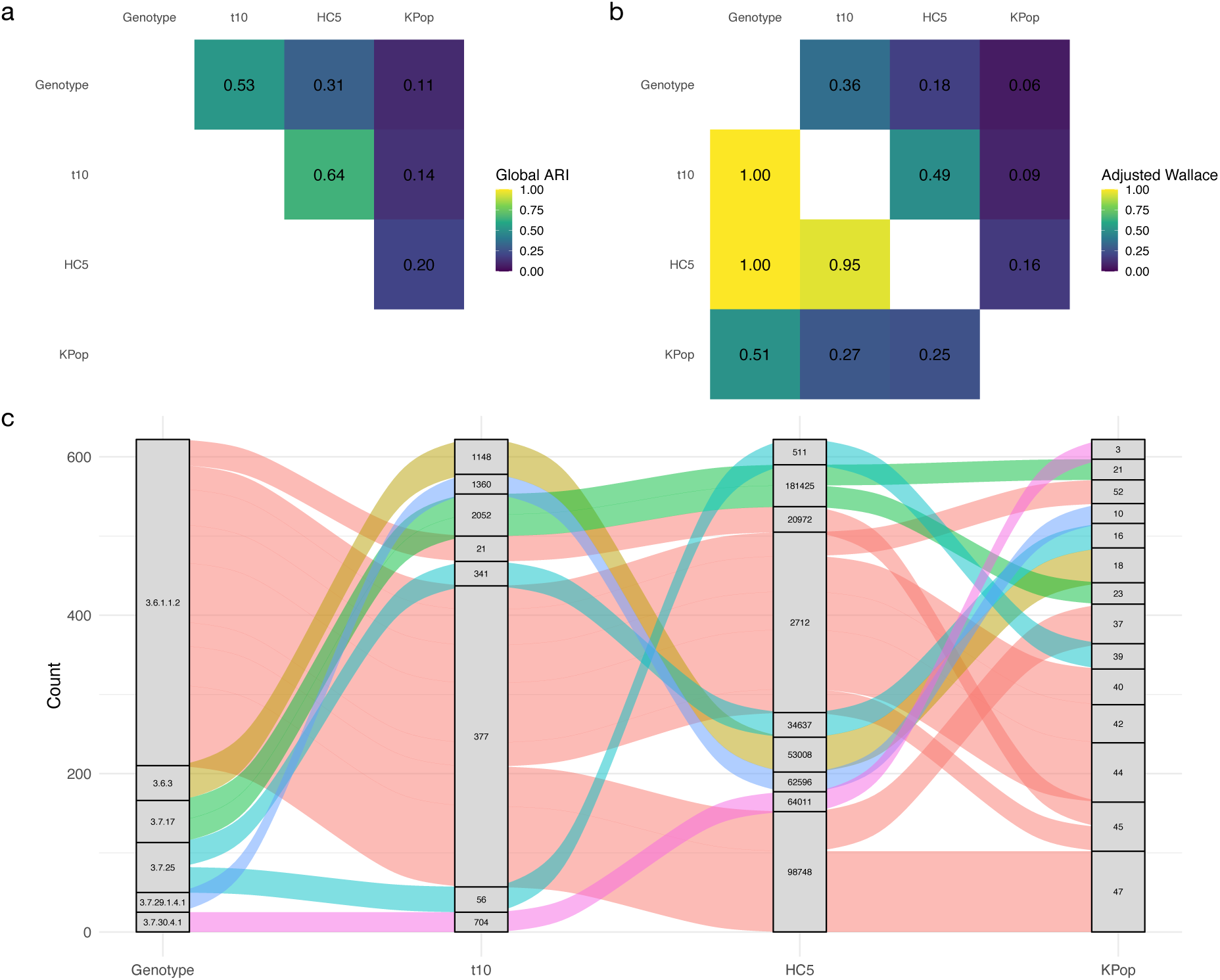
A structured comparison of genomic subtyping scheme results. **(a)** Heatmap showing global Adjusted Rand Index (ARI) between clustering schemes. **(b)** Heatmap showing directional adjusted Wallace (AW) coefficients between clustering schemes. Each cell represents the AW value from the scheme on the y-axis to the scheme on the x-axis, indicating the probability, adjusted for chance, that two isolates grouped together under the row scheme are also grouped together under the column scheme. Higher values reflect stronger predictive correspondence in that direction. **(c)** Alluvial diagram illustrating the 15 most abundant cross-scheme correspondence (flows) between Genotypes, t10 clusters, HC5 clusters, and KPop clusters. Each band (coloured according to Genotype subtype) represents a group of isolates, and its width is proportional to the number of isolates within that group. The flows depict how isolates transition between clustering schema, highlighting structural relationships between methods.

Likewise, AW in the direction Genotype to t10 = 0.361 and Genotype to HC5 = 0.184 were low, whereas the reverse directions were near-deterministic (AW in the direction t10 to Genotype = 0.999; HC5 to Genotype = 0.999), indicating that Genotype was the coarsest scheme. KPop clusters were not nested in either allele-based or SNP-linkage partitions: AW in the direction HC5 to KPop = 0.164 (KPop to HC5 = 0.253) and t10 to KPop = 0.0909 (KPop to t10 = 0.274), indicating an orthogonal community structure derived from k-mer geometry. An alluvial plot from Genotype to t10 to HC5 to KPop (**Figure 3c**) mirrored these AW asymmetries where large t10 flows were split across multiple HC5 and KPop clusters, while HC5 streams mostly remained within single t10 bands. This pattern corresponds to the high AW from HC5 to t10 and the lower AW from t10 to HC5 shown in **Figure 3b**.

### Epidemiological performance of subtyping schema

To assess the performance of subtyping schemes for use in real surveillance of a pathogen with multiple transmission modes, we compared the epidemiological features of clusters subtyped by each method, looking at both demographic composition (i.e. pMSM and non-pMSM) and temporal relationships among isolates.

Regarding the performance of subtyping schema across different transmission modes, we analysed subtypes detected for each demographic group (pMSM versus non-pMSM). The ten largest clusters of each scheme revealed marked differences in size distribution between methods, reflecting the effect of the relative granularity of the methods in the cross-scheme concordance assessment above (**Figure 4a**). Similarly, the distribution of absolute cluster sizes indicated that Genotype generated larger (fewer) clusters on average compared with t10, HC5, and KPop (**Figure 4b**). Genotype, t10 and HC5 produced highly skewed distributions in the pMSM stratum, with one dominant cluster comprising a large proportion of isolates. KPop yielded more evenly distributed cluster sizes and smaller counts per cluster. The dominance of a large cluster was less pronounced in the non-pMSM stratum across all schemes (**Figure 4a-b**), reflecting that this corresponds with a known MSM-associated lineage Genotype 3.6.1.1.2/t10 377 (Mason et al., 2023), and highlighting the problem of employing any single linkage subtyping scheme to detect outbreaks in dense person-to-person transmission scenarios.

**Figure 4:**
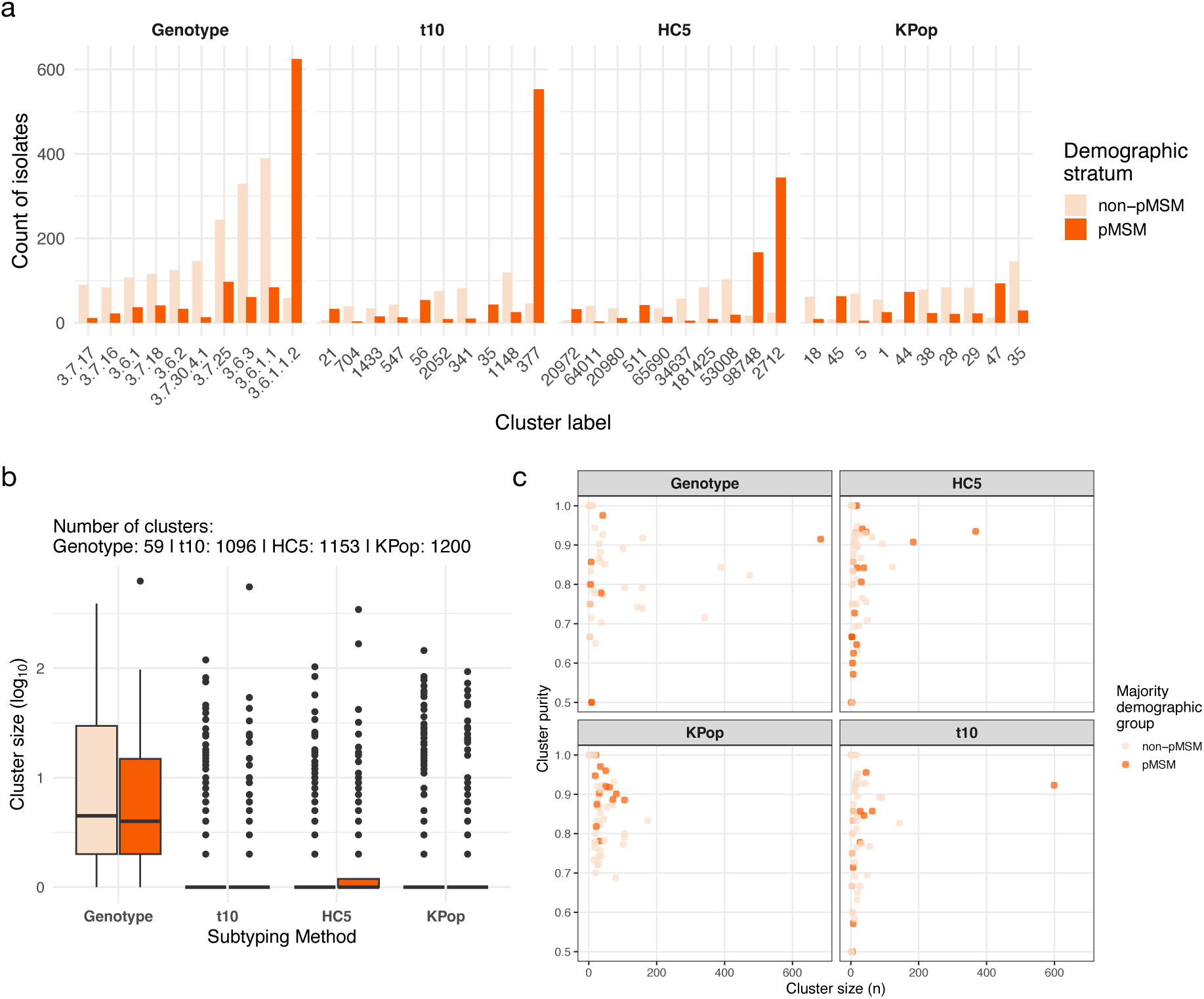
Comparing subtyping schema performance in relation to two major transmission demographics. **(a)** Top ten clusters identified by four clustering methods, stratified by demographic group. Each panel shows the frequency of isolate counts per cluster, stratified by demographic group (coloured according to the inlaid key), for the ten largest clusters for each scheme according to Genotype, t10, HC5, and KPop. **(b)** Distribution of cluster frequencies by genotyping method and stratified by demographic group. Boxplots show the log_10_-transformed frequency of isolates per cluster for each method. Horizontal lines indicate medians, box and whiskers represent the interquartile ranges, and points represent outliers. **(c)** Cluster size (n) and purity across subtyping schemes. Scatter plots of cluster size and purity with respect to demographic grouping (pMSM/non-pMSM) across four subtyping schemes. The dots are coloured by the demographic group that constitutes the majority of isolates within each cluster. Cluster purity is defined as the proportion of isolates in a cluster belonging to the majority demographic group.

To assess the utility of the subtyping schemes for detecting smaller outbreaks across different demographics, we summarised cluster sizes against demographic ‘purity’ i.e. proportion of isolates belonging to single demographic (either pMSM or non-pMSM) (**Figure 4c**). Both t10 and HC5 achieved median purity = 1.00 against the demographic, but HC5 distributed isolates more evenly across a larger number of clusters (largest-cluster share = 0.111; 1,153 clusters) compared with t10 (0.180; 1,096 clusters). KPop also reached median purity = 1.00, generating the most fine-grained partition (1,200 clusters) and the most even distribution (largest-cluster share = 0.052). Genotype was the coarsest scheme (median purity = 0.857; largest-cluster share = 0.206; 59 clusters). KPop appeared to generate clusters with a high demographic purity, but this may also reflect the, on average, smaller cluster size (**Figure 4b-c**).

To evaluate the temporal coherence of different genotyping schemes, we examined whether isolates sampled closely together in time (i.e. those more likely to represent direct transmissions) were more likely to be assigned to the same cluster than expected by chance (**Figure 5a**). Across all schemes, pairs collected within short time intervals exhibited strong enrichment for co-clustering (i.e. Odds Ratio > 1 for belonging to the same, rather than different schemes), indicating that all methods capture recent transmission or outbreak-related structure. As anticipated, this enrichment declined progressively with increasing temporal separation, consistent with increased evolutionary distance over time such that eventually (i.e. when sampled between ∼100 to 550 days apart depending on the scheme), isolates were equally likely to be assigned to different clusters (i.e. Odds Ratio = 1, **Figure 5a**) for a given inter-isolate sampling time. However, the magnitude and persistence of this signal varied by scheme where HC5 showed the strongest short-term enrichment and longest maintenance of cluster enrichment across time (about 550 days). Encouragingly, the HC5 schema also outperformed the other schemes in assigning different clusters (i.e. Odds Ratio << 1) to the most temporally distant isolates (i.e. those nearer 1000 days apart).

**Figure 5:**
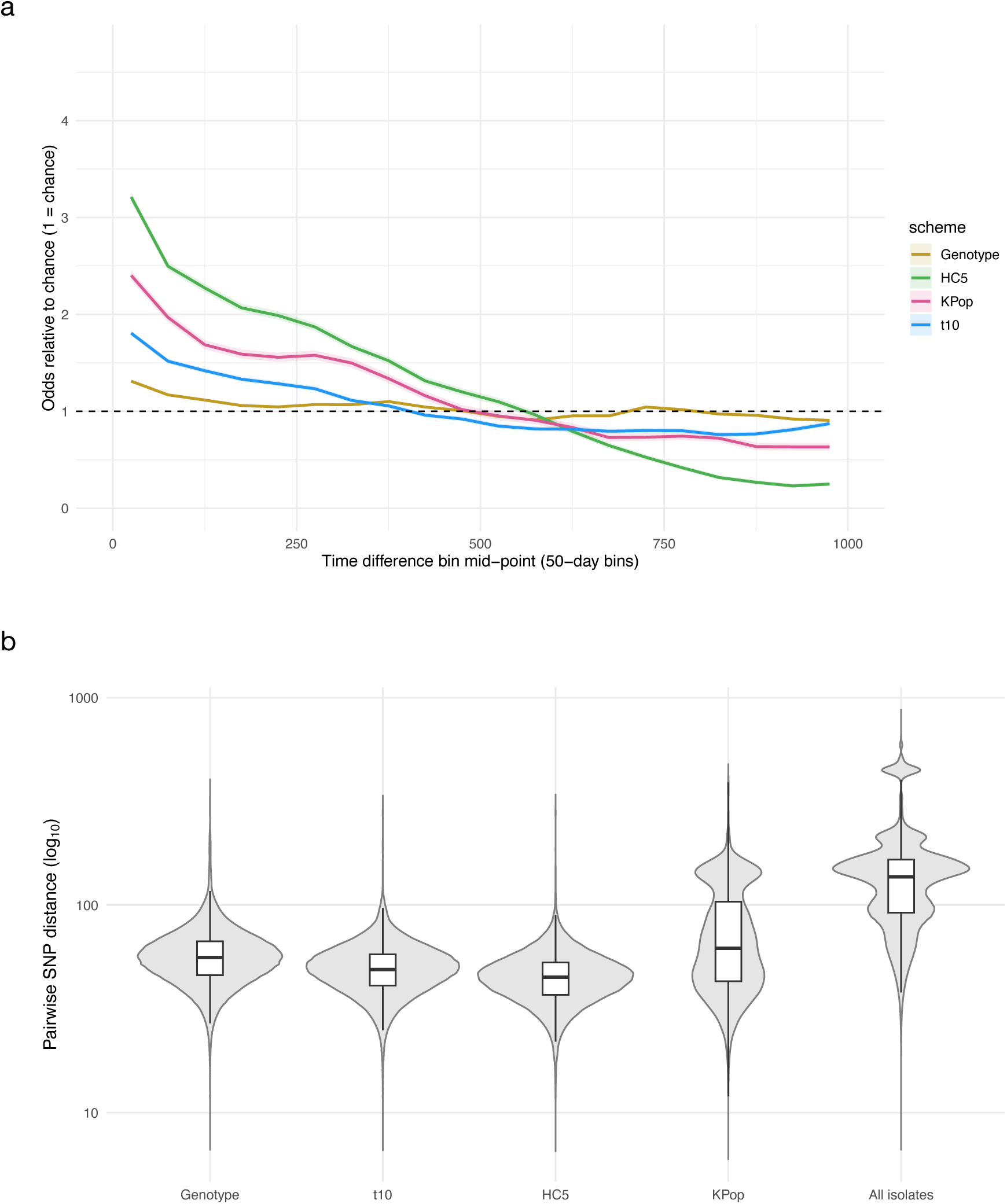
Comparing temporal signal and genetic compactness between and within subtyping schemes. **(a)** Temporal enrichment of clustering schemes. Plot shows odds of two samples being assigned to the same cluster relative to chance (odds ratio = 1) as a function of the time difference between sample collection dates (binned in 50-day intervals, up to 1,000 days). The shaded bands are the exact binomial Clopper-Pearson confidence intervals for each subtyping schemes. **(b)** Violin, with overlaid box and whisker plots, show the distribution of within-cluster pairwise SNP distances from the core genome SNP phylogeny across subtyping schemes and the distribution of pairwise SNP distances of all isolates.

To evaluate the genetic compactness of each subtyping methods, we constructed a maximum likelihood core genome SNP phylogeny of all isolates (**Supplementary Figure 5**) and calculated pairwise patristic distances from the phylogeny among the samples within each cluster. All schemes showed a wide distribution of phylogenetic SNP distances, spanning multiple orders of magnitude, with long upper tails suggesting some isolate pairs inside a cluster were extremely divergent (**Figure 5b**). Overall, in comparing within-cluster phylogenetic distances among Genotype, t10, and HC5, medians reflected the established granularity or the schemes with HC5 having the lowest median pairwise distance. KPop however, had a broader interquartile range and a much higher within-cluster pairwise phylogenetic SNP distance, demonstrating the skew being introduced by a k-mer method which incorporates the bacterial accessory genome.

To explore whether the disconcordant KPop clusterings were attributable to fluctuations in accessory distance, we reviewed the assignments of KPop clusters on the 3.6.1.1.2 Genotype clade on the phylogeny as this clade has undergone several well-described and major accessory genome shifts, (e.g. the acquisition of large ∼100 kb) resistance plasmids (Mason et al., 2023). KPop successfully classified the isolates into two major clusters, reflecting this difference, though many closely related isolates were not assigned to similar KPop clusters relative to HC5 which more clearly captured shifts in both the core and accessory genome (**Supplementary Figure 6**). Hence, although the accessory distances are likely to contribute to meaningful variation in k-mer clustering, our results do not support that this method represents a viable alternative for differentiating *S. sonnei* strains circulating among pMSM based on major changes in the accessory genome.

## Discussion

We compared four genotyping subtyping approaches for *Shigella sonnei* using a large national surveillance dataset and evaluated their epidemiological performance with respect to detecting transmission in different scenarios (sexual vs. non-sexual) and temporal coherence.

Cross-scheme concordance of methods currently in use (i.e. Genotyping, HC5 cgMLST, and t10 SNP) was moderate at the global level, but directional summaries revealed information about the relative granularity of schema. Consistent with previous work, we found that the Genotyping scheme was comparatively coarse relative to HC5 (**Figure 3**) (Hawkey et al., 2021) but even HC5 clustering was unable to sufficiently discriminate large internationally disseminated genomic subtypes resulting from person-to-person transmission in sexual transmission networks (**Figure 4**) (Yassine et al., 2022). Our results also uniquely show that the HC5 clustering offered more granular clustering than the currently employed standard of t10, highlighting the need to move on from these methods, particularly for the discrimination of isolates within large closely related portions of the population structure arising from dense person to person transmission.

When comparing genotyping schemes however, a frequent misconception is to treat granularity as a universal ranking (e.g., whether/how often one scheme splits another’s largest cluster). Our results indicate that resolution depends on the question the scheme is designed to answer. HC5 applies a fixed-radius allele-difference criterion anchored to a portable nomenclature; by construction, it does not merge genomes once they exceed the chosen radius. SNP t10 in contrast, is a single linkage clustering on SNP distances where genomes are aggregated if a path exists through successive ≤ 10-SNP steps. In dense, fast-growing pMSM clades, single linkage clustering tends to percolate through short paths, forming larger aggregates than HC5 would permit, whereas HC5 fragments the same region into multiple groups (reflected in **Figure 4**). Neither behaviour is intrinsically ‘correct’ they simply reflect different operational choices, and there is no ‘ground truth’ with which to formally determine the best performing method.

Distinct from previous work, here we also evaluated the use of k-mer based approaches as tools for *S. sonnei* surveillance and found some promise for the future application of these methods. Although default PopPUNK models under-partitioned the densest, low-diversity region characteristic of pMSM expansions, we applied HDBSCAN (and Leiden community detection, see supplementary) to construct clusters from k-mer distances produced from KPop, which held a greater granularity than HC5 and avoided the over-aggregation typical of single linkage clustering (**Figure 3-4**). These k-mer derived clusters were not anticipated, nor found, to be highly concordant with HC5 or SNP t10 groups (**Figure 3**) potentially as they integrate accessory-genome signals that allele/SNP schemes down-weight or omit. Our results indicate that the value of k-mer based methods is orthogonal and we suggest that they would be most useful for discriminating otherwise poorly discriminated temporally clustered cases, as in genuinely near-clonal segments. All schemes will face a ceiling on discriminability and k-mer based clusters may have the potential to flag rapid shifts in accessory content (e.g., acquisition or replacement of mobile elements). This would be an important advance for sexually transmissible shigellosis, where AMR plasmids and mobile elements are known to be commonly exchanged within the closely connected transmission network (Baker et al., 2018). However, given the broader phylogenetic distance within k-mer based clusters (**Figure 5**) and the poor cluster assignment we observed in comparison with the core SNP phylogeny (**Supplementary Figure 6**), it is likely that further development is needed. Also importantly, better standards for evaluation across k-mer methods and traditional methods are required, as we have shown the difficulty to draw conclusive arguments about the gain of utility from k-mer methods.

In light of the above issues regarding comparing subtyping schema based on fragmentation along, we additionally evaluated epidemiological consistency with respect to the enrichment of clusters for a specific demographic reflective of distinct transmission networks (i.e. purity of either sexual/pMSM or non-sexual/non-pMSM) and temporal coherence (i.e. the likelihood of isolates closely related in time being assigned to the same cluster). This isolates from pMSM were more likely to exist in large, closely related clusters that were less tractable to genomic subtyping (**Figure 4**). We further show that schemes performed variably in their ability to generate small clusters that faithfully reflected the two primary transmission scenarios (**Figure 4**). Likely resulting from its higher granularity, we found that HC5 outperformed other methods with regard to temporal coherence, being both more likely to assign isolates close together in time to the same cluster and least likely to assign isolates further apart in time to the same cluster (**Figure 5**).

Overall, our observations support a pragmatic, hybrid surveillance workflow. First, adopt a stable backbone for communication and cross-site harmonisation, using cgMLST/HierCC (HC5) for portable naming and consistent reporting (Yassine et al., 2022). And secondly, apply context-aware micro-resolution in dense, low-diversity HC5 clusters (e.g., within expanding 98748, 2712 clusters linked to pMSM). Alternatives could be either tighten SNP thresholds and/or use average or complete linkage on SNP distances to reduce over-merging, overlay k-mer-based subtyping as an early-warning layer to flag compact subclusters and potential accessory-genome shift, or the employment of newer systems such as the Life Identification Number (LIN) codes (Hennart et al., 2022) which have recently been implemented in Enterobase.

There are several limitations to this study. First, genomic methods for typing *S. sonnei* are limited by the clonality of the pathogen (Zhao et al., 2023); when genetic divergence is very low, no purely genetic method can accurately mirror epidemiological groupings, so the genomic subtyping should always be complemented with epidemiological metadata. Second, we relied on inferred demographic groups and temporal relationships, as proxies for epidemiology without direct exposure links (e.g., clinic, venue, or household data) impacting our purity estimates and reflecting the lack of a ‘gold standard’ for evaluation of performance. Third, SNP-based analyses may have been affected by our choice of reference and recombination masking which could have affected phylogenetic distance inferences. Besides, cgMLST/HierCC choices (scheme version, missing-data handling) and k-mer parameters (choice of k and other parameters, sketching, and graph construction) influence partitions. However, we selected settings based on stability, though feel other choices would likely only yield small differences. Finally, while we interpret clusters with reference to temporality, an explicit time-series analysis was not fully undertaken here.

Future work should include such prospective simulations that stream isolates by collection date to quantify time-to-detection, early purity, and merge rates across schemes. Also, plasmid-aware distances that separate plasmid flux from clonal core signal (e.g., dual-channel distances or down-weighting high-mobility loci) should be explored. Stability diagnostics for operational thresholds (plateaus across resolution parameters, bootstrap concordance bands), and harmonisation artefacts, including how sampling and reporting cadence influence perceived cluster growth in pMSM, should also be examined. Where feasible, integrating epidemiological linkage (clinic, venue, or household) would allow direct estimation of positive predictive value and detection lag, providing the most policy-relevant evaluation.

## Methods

### Study design and isolate inclusion

We analysed *Shigella sonnei* genomes generated through routine national surveillance by UKHSA between January 2016 and December 2022. Isolates with available short-read WGS (platform: Illumina) and essential metadata (collection date, sample identifiers, and minimal epidemiological data) were eligible. We defined presumptive MSM (pMSM) cases based on demographic data combination of male, adult (aged 16 years and above), and without recorded travel history to higher-risk countries (those within Asia, Africa, and Latin America).

### Sequencing data processing and quality control

Raw reads were adapter/quality trimmed using fastp (v0.23.4, Chen et al., 2018). QC metrices were generated using bacQC (v2.0.0, avantonder, 2023) and assemblyqc (v2.2.0, Rashid et al., 2024). Exclusions were applied sequentially as follows (**Figure 1a**). At the read level, eight isolates were removed due to faulty FASTQ files (sequence–quality string length mismatch, n = 3; truncated files, n = 5). After adapter/quality trimming, samples failing any of the following thresholds were excluded — post-trim total bases > 50 Mbp (n = 9), depth of coverage ≥ 20× (n = 64), total reads > 1 million (n = 58), read mean ≥ 90 (n = 11), and average Phred quality ≥ 30 (n = 0) — yielding 66 unique exclusions after deduplication across criteria. A further 97 isolates were removed for retaining < 50% of reads after trimming (139 total failures, of which 42 had already been excluded by the preceding read-QC filters), leaving 3,679 isolates. Species-composition filtering with Kraken2/Bracken (requiring ≤ 5% non-target taxa) removed 38 additional isolates (40 total failures, 2 previously excluded), leaving 3,641 isolates. Reads were assembled using shovill (v1.1.0, Seemann, 2020). Assembly-level QC then excluded 49 isolates that fell outside the acceptance ranges — N50 between 500 and 60,000 bp (n = 7), contig count between 200 and 1,600 (n = 25), and total assembly size < 6.0 Mb (n = 17) — leaving 3,592 genomes. Finally, 56 isolates lacking essential outputs (SNP address and/or cgMLST and/or *in silico S. sonnei* genotyping by sonneityping [v20210201; Hawkey et al., 2021]) and two identified as non-*sonnei* by sonneityping were removed, yielding the final analytical set of 3,534 genomes. The retained assemblies had a median total assembly size of 4.67 Mb (IQR: 4.60-4.77 Mb), with contig N50 length 23 kb (IQR: 22-23 kb) and L50 count 61 (IQR: 59-62). Assemblies comprised a median of 772 (IQR: 678-884) contigs. The GC content was 50.6% (IQR: 50.5-50.6%), and there were no ambiguous bases (N).

### Antimicrobial resistance markers

AMR determinants were identified from reads/assemblies with AMRFinderPlus (v3.12.8, Feldgarden et al., 2021). Azithromycin (AZM) resistance is defined by presence of genes *mph(A)* and/or *erm(B)*. Ciprofloxacin (CIP) resistance is defined by the presence of at least three quinolone resistance-determining region (QRDR) mutations (within genes *gyrA* and/or *parC*). Ceftriaxone (CRO) resistance is defined by the presence of at least one *bla*CTX-M gene (Mason et al., 2023).

### Subtyping cluster generation

Hierarchical single-linkage clustering was applied to generate SNP-address codes with UKHSA’s in-house implementation of SnapperDB (Dallman et al., 2018). cgMLST allele profiles were assigned using EnteroBase (Mellmann et al., 2011). Hierarchical clustering levels (Zhou et al., 2021) were extracted from EnteroBase to provide comparable radii in allele-difference space. *S. sonnei* genotype labels (e.g., 3.x.y.z) were assigned with an *in silico* sonnei-genotyping script (v20210201; Hawkey et al., 2021) embedded in Mykrobe (v0.13.0; Bradley et al., 2015).

PopPUNK (v2.7.6; Lees et al., 2019) was used to generate whole-genome k-mer sketches and fit strain-level clustering models for all *S. sonnei* isolates. The full dataset was initially analysed as a single set, then separate input lists were created for pMSM and non-pMSM strata. Databases were built with default sketch parameters and varying k-mer sizes. Model fitting was first attempted using DBSCAN, followed by Gaussian mixture modelling (BGMM) to evaluate alternative cluster boundaries. Lineage mode (Zhao et al., 2023) was additionally run to assess broader genomic structure.

KPop (Didelot and Ribeca, 2025) computes full k-mer spectra from each sequence assembly and learns a dataset-specific Correspondence Analysis (CA) transformation (“twister”) to produce low-dimensional embeddings. We ran kpop-workflow (v1.0.0, Morrison and Ribeca, 2025) under cluster mode, and restricted maximum dimension to 100. We then used the resulting pairwise χ^2^/CA distance matrix for clustering. We applied HDBSCAN and carried out a parameter sweep over minimal cluster size to evaluate the number of recovered clusters, fraction of points labelled as noise, and mean silhouette coefficient. We decided to set the threshold as 15 because it maximises the mean silhouette coefficient and minimises the proportion of noises; this yielded 52 non-singleton clusters. We analysed the 1,149 noises and found this comparable to the fraction of singletons in t10 and HC5 (**Supplementary Figure 4**). Alternatively, we constructed k-nearest-neighbour (kNN) graphs from KPop distances. Leiden community detection was run across a resolution grid γ ∈ {0.025, 0.05, 0.075, 0.10, 0.25, 0.50, 1.0} and k ∈ {10, 20, 30, 40, 50}. Stable solution at k = 20, γ = 0.05 maximised ARI against other subtyping schemes, yielding 77 clusters.

### Phylogenomic inference

Whole-genome phylogenetic reconstruction was performed using a reference-based SNP-calling workflow implemented through the P-DOR pipeline (v1.1; Batisti Biffignandi et al., 2023). Illumina paired-end reads for all isolates (n = 3,535) were aligned to the *Shigella sonnei* 53G reference genome (NC_016822.1), and core-genome SNPs were identified after filtering low-quality and ambiguous sites. P-DOR generates a core-SNP alignment and an initial maximum-likelihood (ML) phylogeny using IQ-TREE (Nguyen et al., 2015) with the settings “-m MFP+GTR+ASC -bb 1000”. Recombination was subsequently removed using Gubbins (v3; Croucher et al., 2015), with the ML tree from P-DOR supplied as a starting tree (--bootstrap 1000). The resulting recombination-filtered core SNP alignment was then used to infer the final maximum-likelihood phylogeny in IQ-TREE with the same settings. Pairwise SNP distances were also generated by P-DOR and used for complementary comparative analyses. Tip annotations included epidemiological metadata, scheme labels (Genotype, t10, HC5, KPop clusters), demographic strata, and AMR markers. Tree visualisations used iTOL (Letunic and Bork, 2024).

### Concordance and directional predictiveness

Cross-scheme agreement was measured with several pairwise clustering agreement metrics, including Adjusted Rand Index (ARI; *mclust*), Normalised Mutual Information (NMI; *aricode*), Adjusted Mutual Information (AMI), and directional Adjusted Wallace (AW). Directional AW coefficients were computed following the definition of Carriço/Severiano (probability that two items in the same cluster under scheme A are also clustered together under scheme B, adjusted for chance). Because AW is asymmetric, we reported both directions. All metrics were calculated over the full dataset and stratified by demographic group.

To assess temporal signal in co-occurrence, we calculated time-lagged counts of all unordered sample pairs and the subset belonging to the same cluster, for lags from 0 days to the full observation window. For each lag bin, we estimated the probability that two samples separated by that time difference fall in the same cluster, and derived exact Clopper-Pearson binomial 95% confidence intervals. Enrichment was defined as the ratio of this lag-specific probability to the overall same-cluster probability expected from global cluster-size marginals.

### Statistical analyses and visualisations

Two-sided p < 0.05 was considered statistically significant unless otherwise stated. Analyses and visualisations were performed in R (v4.4.2) and Python (v3.12.12).

## List of abbreviations

ARI: Adjusted Rand Index
AMI: Adjusted Mutual Information
AMR: Antimicrobial Resistance
AW: Adjusted Wallace
AZM: Azithromycin
BGMM: Bayesian Gaussian Mixture Models
CA: Correspondence Analysis
CIP: Ciprofloxacin
CRO: Ceftriaxone
HDBSCAN: Hierarchical Density-Based Spatial Clustering of Applications with Noise
kNN: k-nearest-neighbour
LMICs: Low- and Middle-Income Countries
minPts: minimum cluster size (usually in DBSCAN)
ML: maximum-likelihood
MSM: Men who have Sex with Men
NMI: Normalised Mutual Information
pMSM: presumptive Men who have Sex with Men
QC: Quality Control
QRDR: Quinolone resistance-determining region
SNP: Single-Nucleotide Polymorphism
WGS: Whole-Genome Sequencing
XDR: extensively Drug-Resistant

## Declarations

### Ethics approval and consent to participate

No individual patient consent was required or sought as UKHSA has authority to handle patient data for public health monitoring and infection control under section 251 of the UK National Health Service Act of 2006 (previously section 60 of the Health and Social Care Act of 2001).

### Consent for publication

Not applicable

### Clinical trial number

Not applicable

### Availability of data and materials

All quality-controlled *Shigella sonnei* isolates used in this study (SRR number), accompanied with their metadata, are available in **Supplementary Table 3**.

### Authors’ contributions

K.S.B., C.J. and K.C.W. designed the study; K.C.W. undertook analyses; C.E.C., L.C.E.M., G.B.B. supported the analyses; R.M. answered questions about the methodologies; C.J. and K.S.B. supervised the study; K.C.W. wrote the first draft of the manuscript; K.C.W., L.C.E.M, C.J. and K.S.B. edited the manuscript. All authors had access to the data and read, contributed and approved the final manuscript.

## Acknowledgements

One of the authors is employed by the MPAI Store, a not-for-profit organisation whose mission is to distribute and promote AI workflows compliant with MPAI standards. The workflow described in this paper is open source and hosted independently of the MPAI Store. The authors declare that this work was conducted independently and that their employment did not influence the design, implementation, or findings reported herein.

## References

Achtman, M., Zhou, Z., Charlesworth, J., Baxter, L., 2022. EnteroBase: hierarchical clustering of 100 000s of bacterial genomes into species/subspecies and populations. Philos. Trans. R. Soc. B Biol. Sci. 377, 20210240. 10.1098/rstb.2021.0240

Armstrong, G.L., MacCannell, D.R., Carleton, H.A., Neuhaus, E.B., Bradbury, R.S., Posey, J.E., Taylor, J., Gwinn, M., 2019. Pathogen Genomics in Public Health. N. Engl. J. Med. 381, 2569–2580. 10.1056/NEJMsr1813907

avantonder, 2023. avantonder/bacQC.

Baker, D.J., Robbins, A., Newman, J., Anand, M., Wolfgang, W.J., Mendez-Vallellanes, D.V., Wirth, S.E., Mingle, L.A., 2023. Challenges Associated with Investigating Salmonella Enteritidis with Low Genomic Diversity in New York State: The Impact of Adjusting Analytical Methods and Correlation with Epidemiological Data. Foodborne Pathog. Dis. 20, 230–236. 10.1089/fpd.2022.0068

Baker, K.S., Dallman, T.J., Thomson, N.R., Jenkins, C., 2018. An outbreak of a rare Shiga-toxin-producing Escherichia coli serotype (O117:H7) among men who have sex with men. Microb. Genomics 4, e000181. 10.1099/mgen.0.000181

Batisti Biffignandi, G., Bellinzona, G., Petazzoni, G., Sassera, D., Zuccotti, G.V., Bandi, C., Baldanti, F., Comandatore, F., Gaiarsa, S., 2023. P-DOR, an easy-to-use pipeline to reconstruct bacterial outbreaks using genomics. Bioinformatics 39, btad571. 10.1093/bioinformatics/btad571

Bradley, P., Gordon, N.C., Walker, T.M., Dunn, L., Heys, S., Huang, B., Earle, S., Pankhurst, L.J., Anson, L., de Cesare, M., Piazza, P., Votintseva, A.A., Golubchik, T., Wilson, D.J., Wyllie, D.H., Diel, R., Niemann, S., Feuerriegel, S., Kohl, T.A., Ismail, N., Omar, S.V., Smith, E.G., Buck, D., McVean, G., Walker, A.S., Peto, T.E.A., Crook, D.W., Iqbal, Z., 2015. Rapid antibiotic-resistance predictions from genome sequence data for Staphylococcus aureus and Mycobacterium tuberculosis. Nat. Commun. 6, 10063. 10.1038/ncomms10063

Charles, H., Prochazka, M., Thorley, K., Crewdson, A., Greig, D.R., Jenkins, C., Painset, A., Fifer, H., Browning, L., Cabrey, P., Smith, R., Richardson, D., Waters, L., Sinka, K., Godbole, G., Corkin, H., Abrahams, A., LeBlond, H., Lo, J., Holgate, A., Saunders, J., Plahe, G., Vusirikala, A., Green, F., King, M., Tewolde, R., Jajja, A., 2022. Outbreak of sexually transmitted, extensively drug-resistant Shigella sonnei in the UK, 2021–22: a descriptive epidemiological study. Lancet Infect. Dis. 22, 1503–1510. 10.1016/S1473-3099(22)00370-X

Charles, H., Sinka, K., Simms, I., Baker, K.S., Godbole, G., Jenkins, C., 2024. Trends in shigellosis notifications in England, January 2016 to March 2023. Epidemiol. Infect. 152, e115. 10.1017/S0950268824001006

Chen, S., Zhou, Y., Chen, Y., Gu, J., 2018. fastp: an ultra-fast all-in-one FASTQ preprocessor. Bioinformatics 34, i884–i890. 10.1093/bioinformatics/bty560

Croucher, N.J., Page, A.J., Connor, T.R., Delaney, A.J., Keane, J.A., Bentley, S.D., Parkhill, J., Harris, S.R., 2015. Rapid phylogenetic analysis of large samples of recombinant bacterial whole genome sequences using Gubbins. Nucleic Acids Res. 43, e15. 10.1093/nar/gku1196

Dallman, T., Ashton, P., Schafer, U., Jironkin, A., Painset, A., Shaaban, S., Hartman, H., Myers, R., Underwood, A., Jenkins, C., Grant, K., 2018. SnapperDB: a database solution for routine sequencing analysis of bacterial isolates. Bioinformatics 34, 3028–3029. 10.1093/bioinformatics/bty212

Dallman, T.J., Byrne, L., Ashton, P.M., Cowley, L.A., Perry, N.T., Adak, G., Petrovska, L., Ellis, R.J., Elson, R., Underwood, A., Green, J., Hanage, W.P., Jenkins, C., Grant, K., Wain, J., 2015. Whole-Genome Sequencing for National Surveillance of Shiga Toxin–Producing Escherichia coli O157. Clin. Infect. Dis. Off. Publ. Infect. Dis. Soc. Am. 61, 305–312. 10.1093/cid/civ318

den Bakker, H.C., Allard, M.W., Bopp, D., Brown, E.W., Fontana, J., Iqbal, Z., Kinney, A., Limberger, R., Musser, K.A., Shudt, M., Strain, E., Wiedmann, M., Wolfgang, W.J., 2014. Rapid Whole-Genome Sequencing for Surveillance of Salmonella enterica Serovar Enteritidis. Emerg. Infect. Dis. 20, 1306–1314. 10.3201/eid2008.131399

Didelot, X., Ribeca, P., 2025. KPop: accurate and scalable comparative analysis of microbial genomes by sequence embeddings. Genome Biol. 26, 170. 10.1186/s13059-025-03585-8

DuPont, H.L., Levine, M.M., Hornick, R.B., Formal, S.B., 1989. Inoculum Size in Shigellosis and Implications for Expected Mode of Transmission. J. Infect. Dis. 159, 1126–1128. 10.1093/infdis/159.6.1126

ECDC, 2019. ECDC strategic framework for the integration of molecular and genomic typing into European surveillance and multi-country outbreak investigations: 2019–2021. Publications Office, LU.

Feldgarden, M., Brover, V., Gonzalez-Escalona, N., Frye, J.G., Haendiges, J., Haft, D.H., Hoffmann, M., Pettengill, J.B., Prasad, A.B., Tillman, G.E., Tyson, G.H., Klimke, W., 2021. AMRFinderPlus and the Reference Gene Catalog facilitate examination of the genomic links among antimicrobial resistance, stress response, and virulence. Sci. Rep. 11, 12728. 10.1038/s41598-021-91456-0

Gaudreau, C., Bernaquez, I., Pilon, P.A., Goyette, A., Yared, N., Bekal, S., 2022. Clinical and Genomic Investigation of an International Ceftriaxone- and Azithromycin-Resistant Shigella sonnei Cluster among Men Who Have Sex with Men, Montréal, Canada 2017–2019. Microbiol. Spectr. 10, e02337–21. 10.1128/spectrum.02337-21

Hawkey, J., Paranagama, K., Baker, K.S., Bengtsson, R.J., Weill, F.-X., Thomson, N.R., Baker, S., Cerdeira, L., Iqbal, Z., Hunt, M., Ingle, D.J., Dallman, T.J., Jenkins, C., Williamson, D.A., Holt, K.E., 2021. Global population structure and genotyping framework for genomic surveillance of the major dysentery pathogen, Shigella sonnei. Nat. Commun. 12, 2684. 10.1038/s41467-021-22700-4

Hennart, M., Guglielmini, J., Bridel, S., Maiden, M.C.J., Jolley, K.A., Criscuolo, A., Brisse, S., 2022. A Dual Barcoding Approach to Bacterial Strain Nomenclature: Genomic Taxonomy of Klebsiella pneumoniae Strains. Mol. Biol. Evol. 39, msac135. 10.1093/molbev/msac135

Ingle, D.J., Easton, M., Valcanis, M., Seemann, T., Kwong, J.C., Stephens, N., Carter, G.P., Gonçalves da Silva, A., Adamopoulos, J., Baines, S.L., Holt, K.E., Chow, E.P.F., Fairley, C.K., Chen, M.Y., Kirk, M.D., Howden, B.P., Williamson, D.A., 2019. Co-circulation of Multidrug-resistant Shigella Among Men Who Have Sex With Men in Australia. Clin. Infect. Dis. 69, 1535–1544. 10.1093/cid/ciz005

Jolley, K.A., Bray, J.E., Maiden, M.C.J., 2018. Open-access bacterial population genomics: BIGSdb software, the PubMLST.org website and their applications. Wellcome Open Res. 3, 124. 10.12688/wellcomeopenres.14826.1

Katz, L.S., Griswold, T., Williams-Newkirk, A.J., Wagner, D., Petkau, A., Sieffert, C., Van Domselaar, G., Deng, X., Carleton, H.A., 2017. A Comparative Analysis of the Lyve-SET Phylogenomics Pipeline for Genomic Epidemiology of Foodborne Pathogens. Front. Microbiol. 8, 375. 10.3389/fmicb.2017.00375

Khalil, I.A., Troeger, C., Blacker, B.F., Rao, P.C., Brown, A., Atherly, D.E., Brewer, T.G., Engmann, C.M., Houpt, E.R., Kang, G., Kotloff, K.L., Levine, M.M., Luby, S.P., MacLennan, C.A., Pan, W.K., Pavlinac, P.B., Platts-Mills, J.A., Qadri, F., Riddle, M.S., Ryan, E.T., Shoultz, D.A., Steele, A.D., Walson, J.L., Sanders, J.W., Mokdad, A.H., Murray, C.J.L., Hay, S.I., Reiner, R.C., 2018. Morbidity and mortality due to shigella and enterotoxigenic Escherichia coli diarrhoea: the Global Burden of Disease Study 1990–2016. Lancet Infect. Dis. 18, 1229–1240. 10.1016/S1473-3099(18)30475-4

Lees, J.A., Harris, S.R., Tonkin-Hill, G., Gladstone, R.A., Lo, S.W., Weiser, J.N., Corander, J., Bentley, S.D., Croucher, N.J., 2019. Fast and flexible bacterial genomic epidemiology with PopPUNK. Genome Res. 29, 304–316. 10.1101/gr.241455.118

Lefèvre, S., Njamkepo, E., Feldman, S., Ruckly, C., Carle, I., Lejay-Collin, M., Fabre, L., Yassine, I., Frézal, L., Pardos de la Gandara, M., Fontanet, A., Weill, F.-X., 2023. Rapid emergence of extensively drug-resistant Shigella sonnei in France. Nat. Commun. 14, 462. 10.1038/s41467-023-36222-8

Letunic, I., Bork, P., 2024. Interactive Tree of Life (iTOL) v6: recent updates to the phylogenetic tree display and annotation tool. Nucleic Acids Res. 52, W78–W82. 10.1093/nar/gkae268

Maiden, M.C.J., van Rensburg, M.J.J., Bray, J.E., Earle, S.G., Ford, S.A., Jolley, K.A., McCarthy, N.D., 2013. MLST revisited: the gene-by-gene approach to bacterial genomics. Nat. Rev. Microbiol. 11, 728–736. 10.1038/nrmicro3093

Marshall, J., Lefrancq, N., Mason, L.C.E., Jawed, F., Tam, Y.L., Jenkins, C., Salje, H., Baker, K., 2026. The Spread of Sexually Transmissible Drug-Resistant Shigellosis. 10.2139/ssrn.6134908

Mason, L.C.E., Charles, H., Thorley, K., Chong, C.E., De Silva, P.M., Jenkins, C., Baker, K.S., 2024. The re-emergence of sexually transmissible multidrug resistant Shigella flexneri 3a, England, United Kingdom. Npj Antimicrob. Resist. 2, 20. 10.1038/s44259-024-00038-3

Mason, L.C.E., Greig, D.R., Cowley, L.A., Partridge, S.R., Martinez, E., Blackwell, G.A., Chong, C.E., De Silva, P.M., Bengtsson, R.J., Draper, J.L., Ginn, A.N., Sandaradura, I., Sim, E.M., Iredell, J.R., Sintchenko, V., Ingle, D.J., Howden, B.P., Lefèvre, S., Njamkepo, E., Weill, F.-X., Ceyssens, P.-J., Jenkins, C., Baker, K.S., 2023. The evolution and international spread of extensively drug resistant Shigella sonnei. Nat. Commun. 14, 1983. 10.1038/s41467-023-37672-w

McInerney, J.O., McNally, A., O’Connell, M.J., 2017. Why prokaryotes have pangenomes. Nat. Microbiol. 2, 17040. 10.1038/nmicrobiol.2017.40

Mellmann, A., Harmsen, D., Cummings, C.A., Zentz, E.B., Leopold, S.R., Rico, A., Prior, K., Szczepanowski, R., Ji, Y., Zhang, W., McLaughlin, S.F., Henkhaus, J.K., Leopold, B., Bielaszewska, M., Prager, R., Brzoska, P.M., Moore, R.L., Guenther, S., Rothberg, J.M., Karch, H., 2011. Prospective Genomic Characterization of the German Enterohemorrhagic Escherichia coli O104:H4 Outbreak by Rapid Next Generation Sequencing Technology. PLOS ONE 6, e22751. 10.1371/journal.pone.0022751

Mitchell, H.D., Mikhail, A.F.W., Painset, A., Dallman, T.J., Jenkins, C., Thomson, N.R., Field, N., Hughes, G., 2019. Use of whole-genome sequencing to identify clusters of Shigella flexneri associated with sexual transmission in men who have sex with men in England: a validation study using linked behavioural data. Microb. Genomics 5, e000311. 10.1099/mgen.0.000311

Mitchell, H.D., Thomson, N.R., Jenkins, C., Dallman, T.J., Painset, A., Kirwan, P., Delpech, V., Mikhail, A.F.W., Field, N., Hughes, G., 2021. Linkage of Whole Genome Sequencing, Epidemiological, and Clinical Data to Understand the Genetic Diversity and Clinical Outcomes of Shigella flexneri among Men Who Have Sex with Men in England. Microbiol. Spectr. 9, e0121321. 10.1128/Spectrum.01213-21

Mixão, V., Pinto, M., Brendebach, H., Sobral, D., Dourado Santos, J., Radomski, N., Majgaard Uldall, A.S., Bomba, A., Pietsch, M., Bucciacchio, A., de Ruvo, A., Castelli, P., Iwan, E., Simon, S., Coipan, C.E., Linde, J., Petrovska, L., Kaas, R.S., Grimstrup Joensen, K., Holtsmark Nielsen, S., Kiil, K., Lagesen, K., Di Pasquale, A., Gomes, J.P., Deneke, C., Tausch, S.H., Borges, V., 2025. Multi-country and intersectoral assessment of cluster congruence between pipelines for genomics surveillance of foodborne pathogens. Nat. Commun. 16, 3961. 10.1038/s41467-025-59246-8

Moore, M.P., Laager, M., Ribeca, P., Didelot, X., 2024. KmerAperture: Retaining k-mer synteny for alignment-free extraction of core and accessory differences between bacterial genomes. PLOS Genet. 20, e1011184. 10.1371/journal.pgen.1011184

Morgan, O., Crook, P., Cheasty, T., Jiggle, B., Giraudon, I., Hughes, H., Jones, S.-M., Maguire, H., 2006. Shigella sonnei Outbreak among Homosexual Men, London. Emerg. Infect. Dis. 12, 1458–1460. 10.3201/eid1209.060282

Morrison, R., Ribeca, P., 2025. ryanmorrison22/kpop-workflow.

Nadon, C., Van Walle, I., Gerner-Smidt, P., Campos, J., Chinen, I., Concepcion-Acevedo, J., Gilpin, B., Smith, A.M., Kam, K.M., Perez, E., Trees, E., Kubota, K., Takkinen, J., Nielsen, E.M., Carleton, H., 2017. PulseNet International: Vision for the implementation of whole genome sequencing (WGS) for global food-borne disease surveillance. Eurosurveillance 22, 30544. 10.2807/1560-7917.ES.2017.22.23.30544

Nguyen, L.-T., Schmidt, H.A., von Haeseler, A., Minh, B.Q., 2015. IQ-TREE: A Fast and Effective Stochastic Algorithm for Estimating Maximum-Likelihood Phylogenies. Mol. Biol. Evol. 32, 268–274. 10.1093/molbev/msu300

Niyogi, S.K., 2005. Shigellosis. J. Microbiol. 43, 133–143.

Rashid, U., Wu, C., Shiller, J., Smith, K., Crowhurst, R., Davy, M., Chen, T.-H., Carvajal, I., Bailey, S., Thomson, S., Deng, C.H., 2024. AssemblyQC: a Nextflow pipeline for reproducible reporting of assembly quality. Bioinformatics 40, btae477. 10.1093/bioinformatics/btae477

Roberts, M.D., Davis, O., Josephs, E.B., Williamson, R.J., 2025. K-mer-based Approaches to Bridging Pangenomics and Population Genetics. Mol. Biol. Evol. 42, msaf047. 10.1093/molbev/msaf047

Rozday, T.J., 2025. Metagenomics distilled: new k-mer-based methods. Nat. Rev. Microbiol. 23, 473–473. 10.1038/s41579-025-01192-9

Scott, T.A., Baker, K.S., Trotter, C., Jenkins, C., Mostowy, S., Hawkey, J., Schmidt, H., Holt, K.E., Thomson, N.R., Baker, S., 2025. Shigella sonnei: epidemiology, evolution, pathogenesis, resistance and host interactions. Nat. Rev. Microbiol. 23, 303–317. 10.1038/s41579-024-01126-x

Seemann, T., 2020. tseemann/shovill.

Shi, G., Dai, Y., Zhou, D., Chen, M., Zhang, J., Bi, Y., Liu, S., Wu, Q., 2025. An alignment-and reference-free strategy using k-mer present pattern for population genomic analyses. Mycology 16, 309–323. 10.1080/21501203.2024.2358868

Siddiq, M., O’Flanagan, H., Richardson, D., Llewellyn, C.D., 2023. Factors associated with sexually transmitted shigella in men who have sex with men: a systematic review. Sex. Transm. Infect. 99, 58–63. 10.1136/sextrans-2022-055583

Tansarli, G.S., Long, D.R., Waalkes, A., Bourassa, L.A., Libby, S.J., Penewit, K., Almazan, J., Matsumoto, J., Bryson-Cahn, C., Rietberg, K., Dell, B.M., Hatley, N.V., Salipante, S.J., Fang, F.C., 2023. Genomic Reconstruction and Directed Interventions in a Multidrug-resistant Shigellosis Outbreak in Seattle, Washington: A Genomic Surveillance Study. Lancet Infect. Dis. 23, 740–750. 10.1016/S1473-3099(22)00879-9

Thorley, K., Charles, H., Greig, D.R., Prochazka, M., Mason, L.C.E., Baker, K.S., Godbole, G., Sinka, K., Jenkins, C., 2023. Emergence of extensively drug-resistant and multidrug-resistant *Shigella flexneri* serotype 2a associated with sexual transmission among gay, bisexual, and other men who have sex with men, in England: a descriptive epidemiological study. Lancet Infect. Dis. 23, 732–739. 10.1016/S1473-3099(22)00807-6

UKHSA, 2024. Sexually transmitted Shigella spp. in England: 2016 to 2023 [WWW Document]. GOV.UK. URL https://www.gov.uk/government/publications/non-travel-associated-shigella-infections/sexually-transmitted-shigella-spp-in-england-2016-to-2023 (accessed 8.13.25).

WHO, 2023a. Whole genome sequencing as a tool to strengthen foodborne disease surveillance and response: Module 2. Whole genome sequencing in foodborne disease outbreak investigations. [WWW Document]. URL https://iris.who.int/bitstream/handle/10665/373460/9789240021242-eng.pdf?sequence=1 (accessed 8.13.25).

WHO, 2023b. Whole genome sequencing as a tool to strengthen foodborne disease surveillance and response: Module 3. Whole genome sequencing in foodborne disease routine surveillance. [WWW Document]. URL https://iris.who.int/bitstream/handle/10665/373522/9789240021266-eng.pdf?sequence=1 (accessed 8.13.25).

WHO, 2022. WHO Immunization, Vaccines and Biologicals: Shigella [WWW Document]. URL https://www.who.int/teams/immunization-vaccines-and-biologicals/diseases/shigella (accessed 8.13.25).

WHO, 2018. Whole genome sequencing for foodborne disease surveillance: landscape paper. World Health Organization, Geneva.

Yassine, I., Lefèvre, S., Hansen, E.E., Ruckly, C., Carle, I., Lejay-Collin, M., Fabre, L., Rafei, R., Clermont, D., de la Gandara, M.P., Dabboussi, F., Thomson, N.R., Weill, F.-X., 2022. Population structure analysis and laboratory monitoring of Shigella by core-genome multilocus sequence typing. Nat. Commun. 13, 551. 10.1038/s41467-022-28121-1

Zhao, B., Lees, J.A., Wu, H., Yang, C., Falush, D., 2023. Genealogical inference and more flexible sequence clustering using iterative-PopPUNK. Genome Res. 33, 988–998. 10.1101/gr.277395.122

Zhou, Z., Charlesworth, J., Achtman, M., 2021. HierCC: a multi-level clustering scheme for population assignments based on core genome MLST. Bioinformatics 37, 3645–3646. 10.1093/bioinformatics/btab234

