## Supplementary Material for "Evaluating Genomic Surveillance Methods for *Shigella sonnei* in a High-Income Setting"

**Introduction**

In the main text, we presented KPop clusters generated via HDBSCAN from the KPop pairwise distances (Didelot and Ribeca, 2025). Besides, we explored an alternative clustering method, a graph/community approach that operate on the distance geometry without assuming tree-likeness. The results are presented here.

**Results**

We constructed k-nearest-neighbour (kNN) graphs from KPop distances. A Leiden sweep across *k* ∈ {10, 20, 30, 40, 50} and resolution γ ∈ {0.025, 0.05, 0.075, 0.1, 0.25, 0.5, 0.8} identified stable plateaus (**SM Figure 1**). When optimised to best match the *in silico* genotyping scheme (Hawkey et al., 2021), Adjusted Rand Index ([ARI], with tie-breaking by Adjusted Mutual Information [AMI] and fewer communities) was maximised with *k* = 50 and γ = 0.075, yielding 44 communities. By contrast, agreement with HC5 (Achtman et al., 2022; Zhou et al., 2021) and t10 (Dallman et al., 2018) peaked at *k* = 20 and γ = 0.05 (77 communities), and across the grid community sizes remained moderate (no single giant component). **Figure 2d** in the main text summarises the community size distribution for this solution.

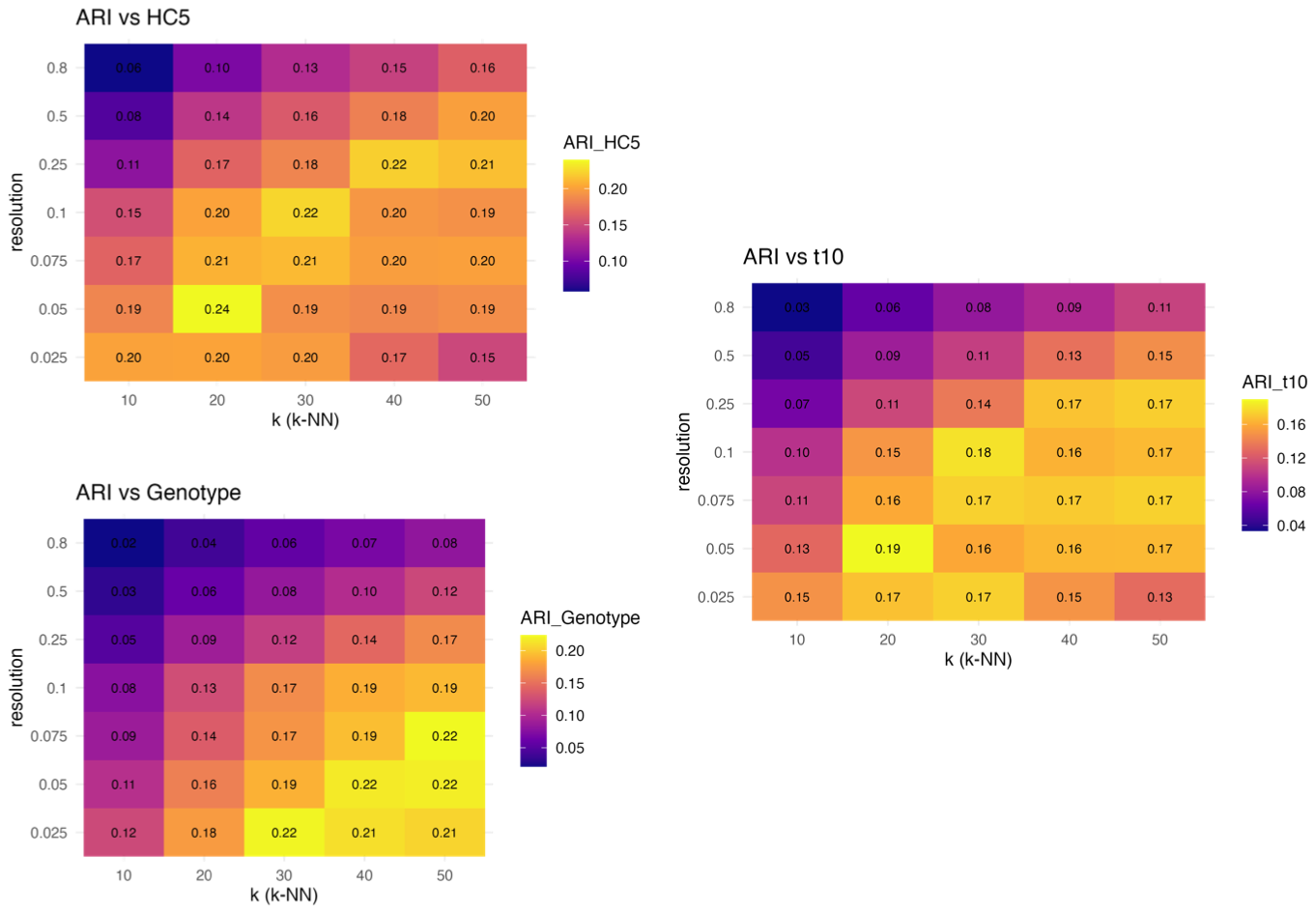

**SM Figure 1**: **Leiden parameter sweep over KPop distances.** ARI values across multiple clustering reference schemes (Genotype, t10, HC5) are shown as a function of the kNN parameter (k = 10-50, resolution = 0.025-0.8).

To assess the concordance between genotyping schemes on this dataset, we compared four schema, Genotyping (Hawkey et al., 2021), cgMLST/HierCC at the HC5 level (Achtman et al., 2022; Zhou et al., 2021), SNP-address at the t10 SNP single linkage cluster level (Dallman et al., 2018), and KPop clusters (derived using Leiden clustering with optimal parameters: k = 20, γ = 0.05, n = 77), using a variety of uni- and bi-directional concordance measures (Adjusted Rand Index [ARI], Normalised Mutual Information [NMI], Adjusted Mutual Information [AMI], and directional Adjusted Wallace [AW]; **SM Table 1**, **SM Figure 2a-b**). An alluvial plot from Genotype to t10 to HC5 to KPop (**SM** **Figure 2c**) mirrored these AW asymmetries where large t10 flows were split across multiple HC5 and KPop clusters, consistent with analyses done with HDBSCAN-derived KPop clusters, while HC5 streams mostly remained within single t10 bands.

**SM Table 1**: **Concordance metrices between subtyping scheme results.**

| **s1** | **s2** | **ARI** | **AMI** | **NMI** | **VI** | **V measure** | **W_s1_to_s2** | **AW_s1_to_s2** | **W_s2_to_s1** | **AW_s2_to_s1** |
| --- | --- | --- | --- | --- | --- | --- | --- | --- | --- | --- |
| Genotype | t10 | 0.5306 | 0.3714 | 0.5164 | 2.6148 | 0.6807 | 0.3856 | 0.3612 | 0.9991 | 0.9989 |
| Genotype | HC5 | 0.3111 | 0.3352 | 0.4895 | 2.9116 | 0.6569 | 0.2005 | 0.1843 | 0.9987 | 0.9985 |
| Genotype | KPop | 0.1591 | 0.3896 | 0.4416 | 3.2739 | 0.5267 | 0.1091 | 0.0928 | 0.6008 | 0.5570 |
| t10 | HC5 | 0.6445 | 0.8173 | 0.9253 | 0.5534 | 0.9501 | 0.4969 | 0.4867 | 0.9555 | 0.9537 |
| t10 | KPop | 0.1897 | 0.3288 | 0.5788 | 3.2741 | 0.6562 | 0.1540 | 0.1385 | 0.3274 | 0.3007 |
| HC5 | KPop | 0.2398 | 0.3217 | 0.5765 | 3.2538 | 0.6687 | 0.2420 | 0.2281 | 0.2675 | 0.2527 |

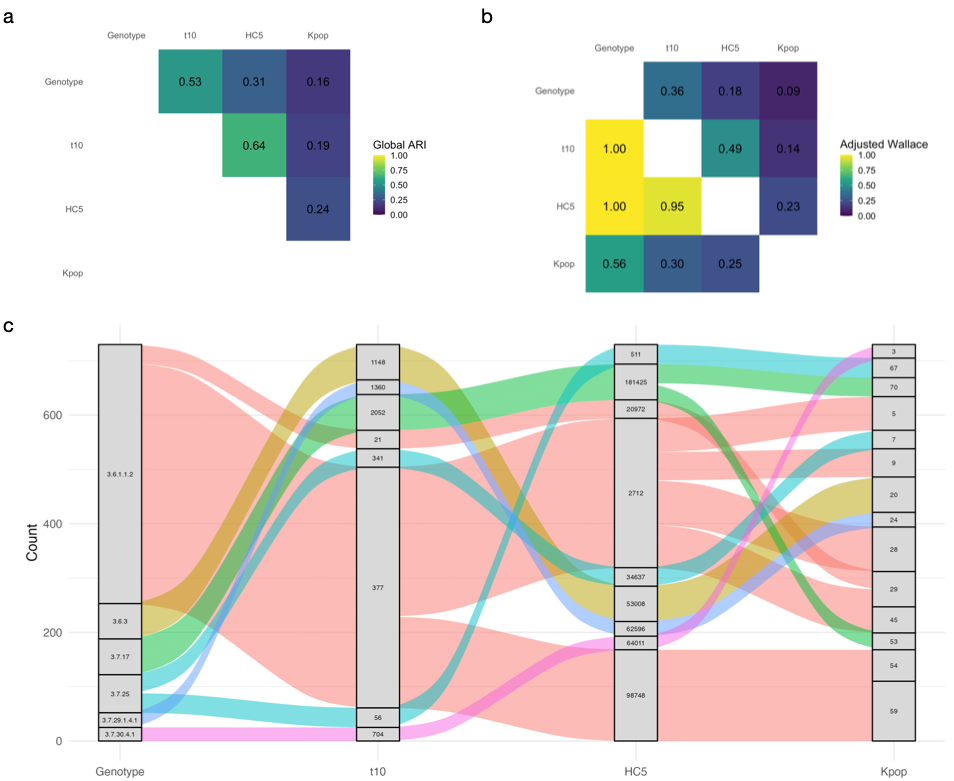

**SM Figure 2**: **A structured** **comparison of genomic subtyping scheme results. (a)** Heatmap showing global Adjusted Rand Index (ARI) between clustering schemes. **(b)** Heatmap showing directional adjusted Wallace (AW) coefficients between clustering schemes. Each cell represents the AW value from the scheme on the y-axis to the scheme on the x-axis, indicating the probability, adjusted for chance, that two isolates grouped together under the row scheme are also grouped together under the column scheme. Higher values reflect stronger predictive correspondence in that direction. **(c)** Alluvial diagram illustrating the 15 most abundant cross-scheme correspondence (flows) between Genotypes, t10 clusters, HC5 clusters, and KPop clusters (derived using Leiden clustering with optimal parameters: k = 20, γ = 0.05, n = 77). Each band (coloured according to Genotype subtype) represents a group of isolates, and its width is proportional to the number of isolates within that group. The flows depict how isolates transition between clustering schema, highlighting structural relationships between methods.

To assess the performance of subtyping schemes for use in real surveillance of a pathogen with multiple transmission modes, we compared the epidemiological features of clusters subtyped by each method, looking at both demographic composition (i.e. pMSM and non-pMSM) and temporal relationships among isolates.

Regarding the performance of subtyping schema across different transmission modes, we analysed subtypes detected for each demographic group (pMSM versus non-pMSM). The ten largest clusters of each scheme revealed marked differences in size distribution between methods, reflecting the effect of the relative granularity of the methods in the cross-scheme concordance assessment above (**SM** **Figure 3a-b**). Similarly, the distribution of absolute cluster sizes indicated that Genotype generated larger (fewer) clusters on average compared with t10, HC5, and KPop (**SM** **Figure 3c**). Consistent with HDBSCAN-derived clusters, KPop (Leiden) yielded more evenly distributed cluster sizes and smaller counts per cluster.

To assess the utility of the subtyping schemes for detecting smaller outbreaks across different demographics, a fragmentation-purity summary (**SM Figure 3d**) showed HC5 achieved median purity = 1.00 against the demographic (pMSM versus non-pMSM) with largest-cluster share = 0.111 across 1,153 clusters, whereas t10 also reached median purity = 1.00 but concentrated more isolates (largest-cluster share = 0.180 across 1,096 clusters). Genotype was coarser (median purity = 0.857; largest share = 0.206; 59 clusters) and KPop spread the signal most evenly (largest share = 0.035 across 77 communities) at the cost of lower behavioural purity (0.84). In contrast to the high purity observed in HDBSCAN-derived KPop clusters (**Figure 4c** in main text), Leiden-derived KPop clusters were more heterogeneous demographically.

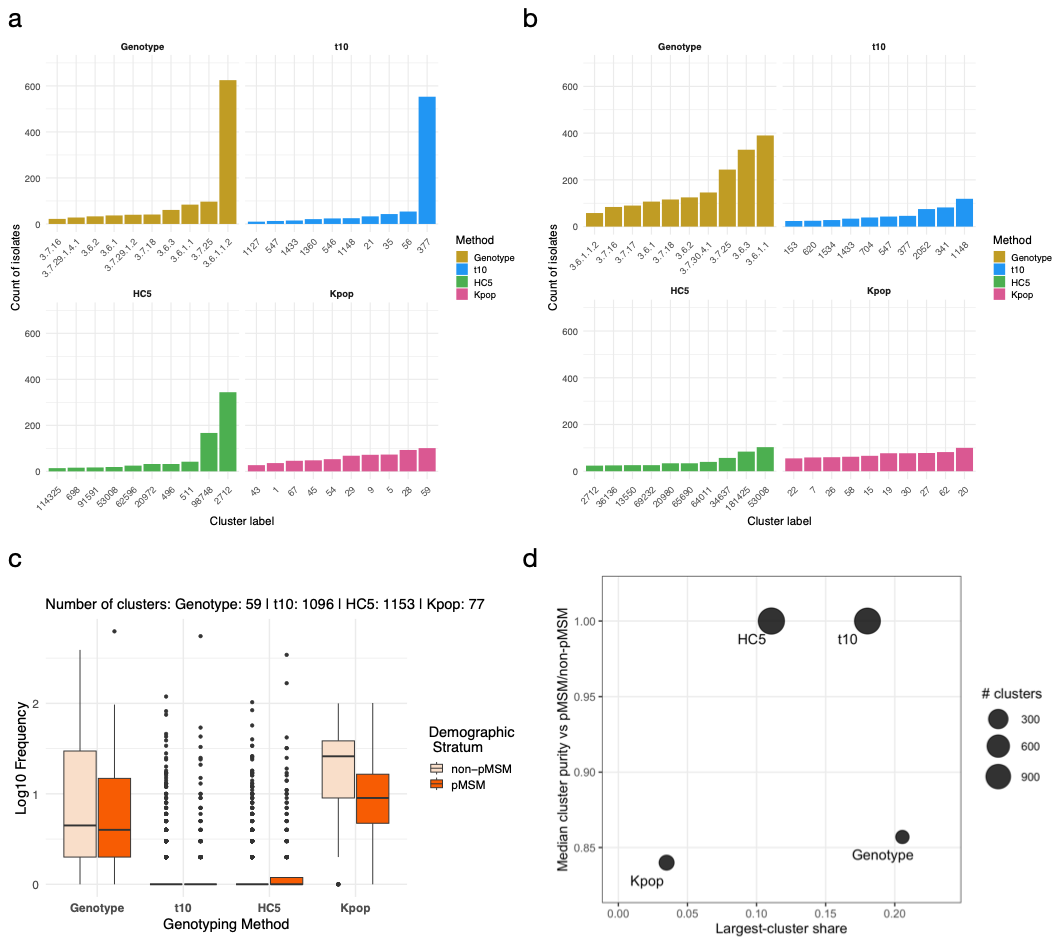

**SM Figure 3**: **Comparing subtyping schema performance in relation to two major transmission demographics. (a)** Top ten clusters identified by four clustering methods in the pMSM demographic group. Each panel shows the frequency of isolate counts per cluster for the ten largest clusters according to genotype, t10, HC5, and KPop. **(b)** Top ten clusters identified by four clustering methods in the non-pMSM demographic group. **(c)** Distribution of cluster frequencies by genotyping method and stratified by demographic group. Boxplots show the log_10_-transformed frequency of isolates per cluster for each method. Orange boxes represent pMSM clusters, and beige boxes represent non-pMSM clusters. Horizontal lines indicate medians, and points represent outliers. **(d)** Fragmentation-purity trade-off. For each scheme, scatterplot of largest-cluster share (x-axis; fraction of isolates in the largest cluster) versus median cluster purity with respect to demographic group (y-axis). Point size is proportional to the number of clusters.

To evaluate the temporal coherence of different genotyping schemes, we examined whether isolates sampled closely together in time (i.e. those more likely to represent direct transmissions) were more likely to be assigned to the same cluster than expected by chance (**SM** **Figure 4**). All schemes showed enrichment for co-clustering at short sampling intervals, with this signal decaying as temporal separation increased. However, the strength and persistence of temporal coherence varied between schemes: HC5 showed the strongest short-term enrichment, maintained cluster coherence for the longest period (~550 days), and most effectively separated the most temporally distant isolates (> 1000 days). Overall, the relative performance of subtyping schemes was consistent with the analyses presented in the main text.

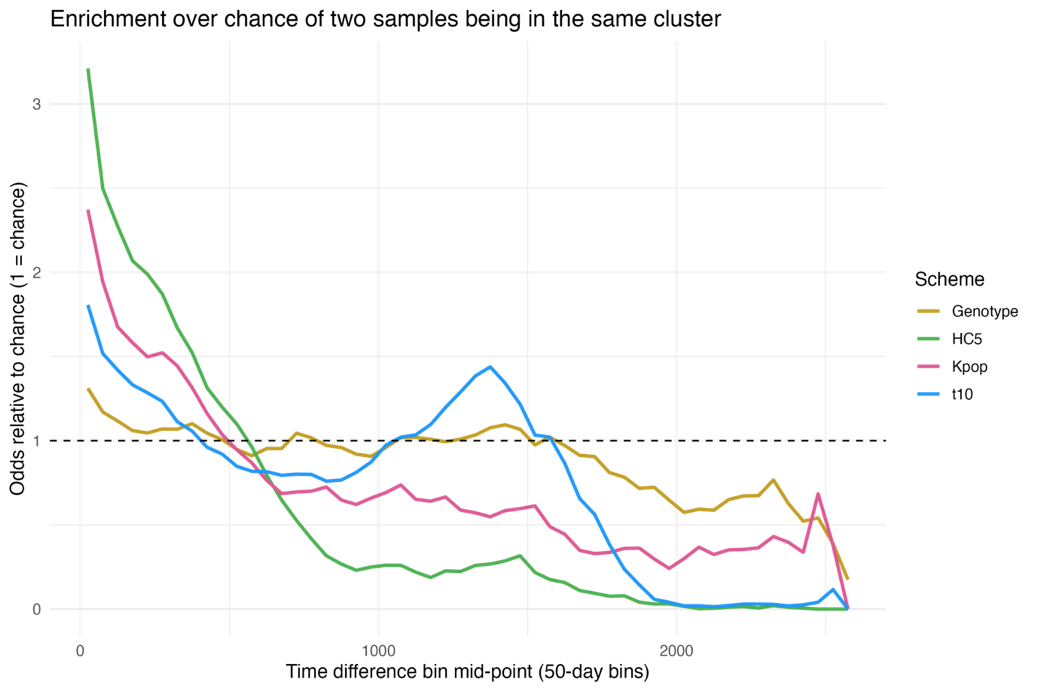

**SM Figure 4**: **Temporal enrichment of clustering schemes.** Plot shows odds of two samples being assigned to the same cluster relative to chance (odds ratio = 1) as a function of the time difference between sample collection dates (binned in 50-day intervals).

To place clustering results in an epidemiological framework, we reconstructed a maximum-likelihood phylogeny of all *S. sonnei* isolates from the dataset (**SM** **Figure 5a**). To explore whether the high within-cluster phylogenetic distances observed for KPop were attributable to closely related accessory distance, we reviewed the assignments of KPop clusters on the 3.6.1.1.2 Genotype clade on the phylogeny (**SM** **Figure 5b**). This clade has undergone an accessory genome shift, leading to a gain of ceftriaxone resistance in 2021-2022 (Mason et al., 2023). KPop successfully identified the latest extensively drug-resistant (XDR) clade as one cluster but classified the earlier clade as more fragmented, smaller clusters. We cannot conclude the utility of incorporating accessory distances by KPop under these results.

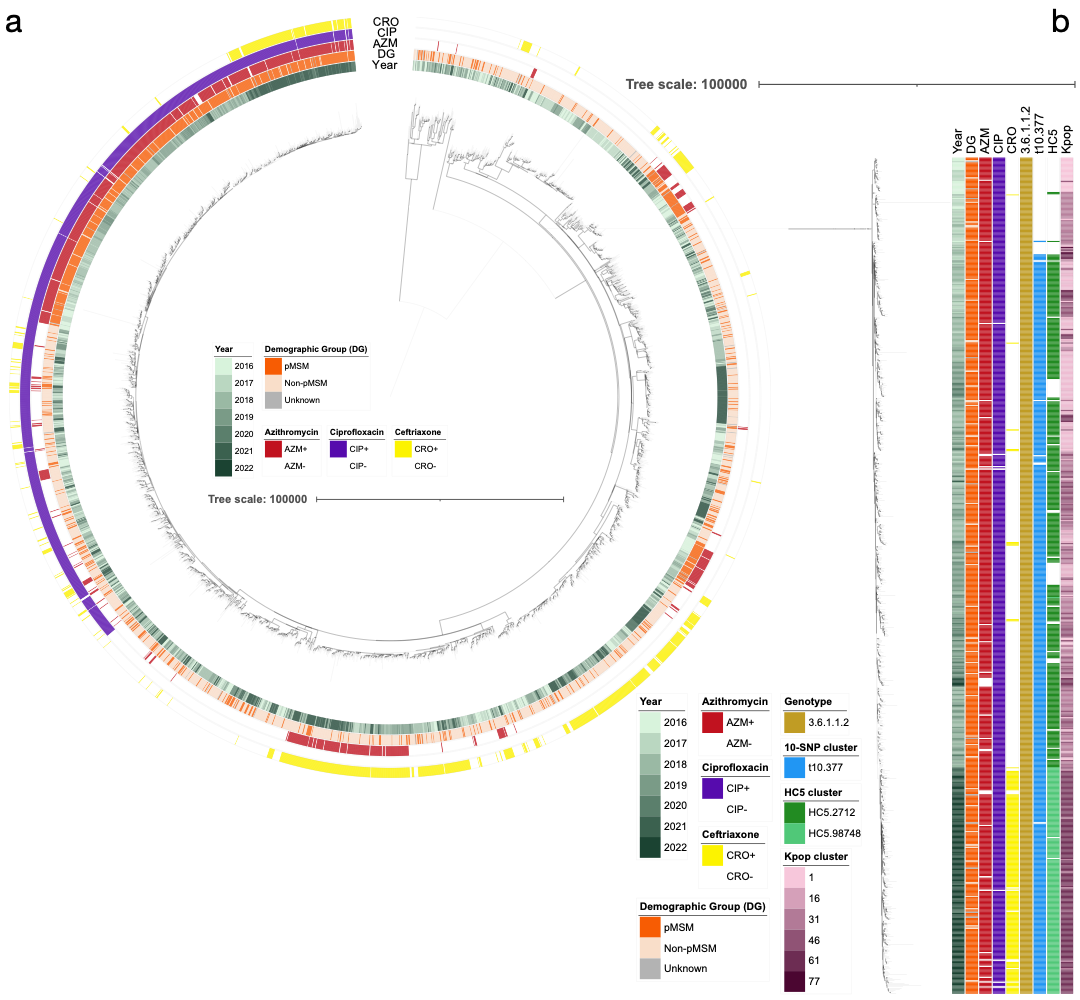

**SM Figure 5**: Maximum-likelihood phylogenetic tree of 3,534 *S. sonnei* isolates from UKHSA routine genomic surveillance. The tree was inferred from a core-SNP alignment (S53G reference) using IQ-TREE (Nguyen et al., 2015) with the best-fit model (MFP + GTR + ASC) and 1,000 bootstrap replicates. Recombination was detected and masked with Gubbins (Croucher et al., 2015) before final tree reconstruction. Branches with support ≥70% are bold. The full tree is shown on the left **(a)** and a focused truncated tree with the terminal 683 nodes is shown on the right **(b)**. Metadata rings: (1) Year of isolation (2016-2022); (2) Demographic group (pMSM, non-pMSM, Unknown); (3) Predicted phenotypic non-susceptibility to azithromycin (AZM), ciprofloxacin (CIP), and ceftriaxone (CRO); (4) Cluster overlays for cross-scheme comparison: (i) *In silico* genotype (sonneityping): example genotype 3.6.1.1.2 highlighted (Hawkey et al., 2021); (ii) SNP-address (SnapperDB): example t10 cluster.377 highlighted (Dallman et al., 2018); (iii) cgMLST/HierCC: example HC5 cluster.2712 and cluster.98748 highlighted (Achtman et al., 2022; Zhou et al., 2021); (iv) KPop clusters (derived using Leiden clustering with optimal parameters: k = 20, γ = 0.05, n = 77), the clusters are labelled with a colour gradient. Missing data are rendered as blank segments.

**Conclusion**

In conclusion, KPop clusters derived from Leiden did not offer significant improvement in resolution or temporal coherence, and performed worse in demographic-specific clustering, compared to the clusters derived from HDBSCAN. Fundamentally, Leiden forces too many singletons into major clusters, which dilutes epidemiologically meaningful structure and reduces the ability to distinguish recent, demographically coherent transmission events.

**References**

Achtman, M., Zhou, Z., Charlesworth, J., Baxter, L., 2022. EnteroBase: hierarchical clustering of 100 000s of bacterial genomes into species/subspecies and populations. Philos. Trans. R. Soc. B Biol. Sci. 377, 20210240. https://doi.org/10.1098/rstb.2021.0240

Croucher, N.J., Page, A.J., Connor, T.R., Delaney, A.J., Keane, J.A., Bentley, S.D., Parkhill, J., Harris, S.R., 2015. Rapid phylogenetic analysis of large samples of recombinant bacterial whole genome sequences using Gubbins. Nucleic Acids Res. 43, e15. https://doi.org/10.1093/nar/gku1196

Dallman, T., Ashton, P., Schafer, U., Jironkin, A., Painset, A., Shaaban, S., Hartman, H., Myers, R., Underwood, A., Jenkins, C., Grant, K., 2018. SnapperDB: a database solution for routine sequencing analysis of bacterial isolates. Bioinformatics 34, 3028–3029. https://doi.org/10.1093/bioinformatics/bty212

Didelot, X., Ribeca, P., 2025. KPop: accurate and scalable comparative analysis of microbial genomes by sequence embeddings. Genome Biol. 26, 170. https://doi.org/10.1186/s13059-025-03585-8

Hawkey, J., Paranagama, K., Baker, K.S., Bengtsson, R.J., Weill, F.-X., Thomson, N.R., Baker, S., Cerdeira, L., Iqbal, Z., Hunt, M., Ingle, D.J., Dallman, T.J., Jenkins, C., Williamson, D.A., Holt, K.E., 2021. Global population structure and genotyping framework for genomic surveillance of the major dysentery pathogen, Shigella sonnei. Nat. Commun. 12, 2684. https://doi.org/10.1038/s41467-021-22700-4

Mason, L.C.E., Greig, D.R., Cowley, L.A., Partridge, S.R., Martinez, E., Blackwell, G.A., Chong, C.E., De Silva, P.M., Bengtsson, R.J., Draper, J.L., Ginn, A.N., Sandaradura, I., Sim, E.M., Iredell, J.R., Sintchenko, V., Ingle, D.J., Howden, B.P., Lefèvre, S., Njamkepo, E., Weill, F.-X., Ceyssens, P.-J., Jenkins, C., Baker, K.S., 2023. The evolution and international spread of extensively drug resistant Shigella sonnei. Nat. Commun. 14, 1983. https://doi.org/10.1038/s41467-023-37672-w

Nguyen, L.-T., Schmidt, H.A., von Haeseler, A., Minh, B.Q., 2015. IQ-TREE: A Fast and Effective Stochastic Algorithm for Estimating Maximum-Likelihood Phylogenies. Mol. Biol. Evol. 32, 268–274. https://doi.org/10.1093/molbev/msu300

Zhou, Z., Charlesworth, J., Achtman, M., 2021. HierCC: a multi-level clustering scheme for population assignments based on core genome MLST. Bioinformatics 37, 3645–3646. https://doi.org/10.1093/bioinformatics/btab234
