## Supplementary Figure for "Evaluating Genomic Surveillance Methods for *Shigella sonnei* in a High-Income Setting"

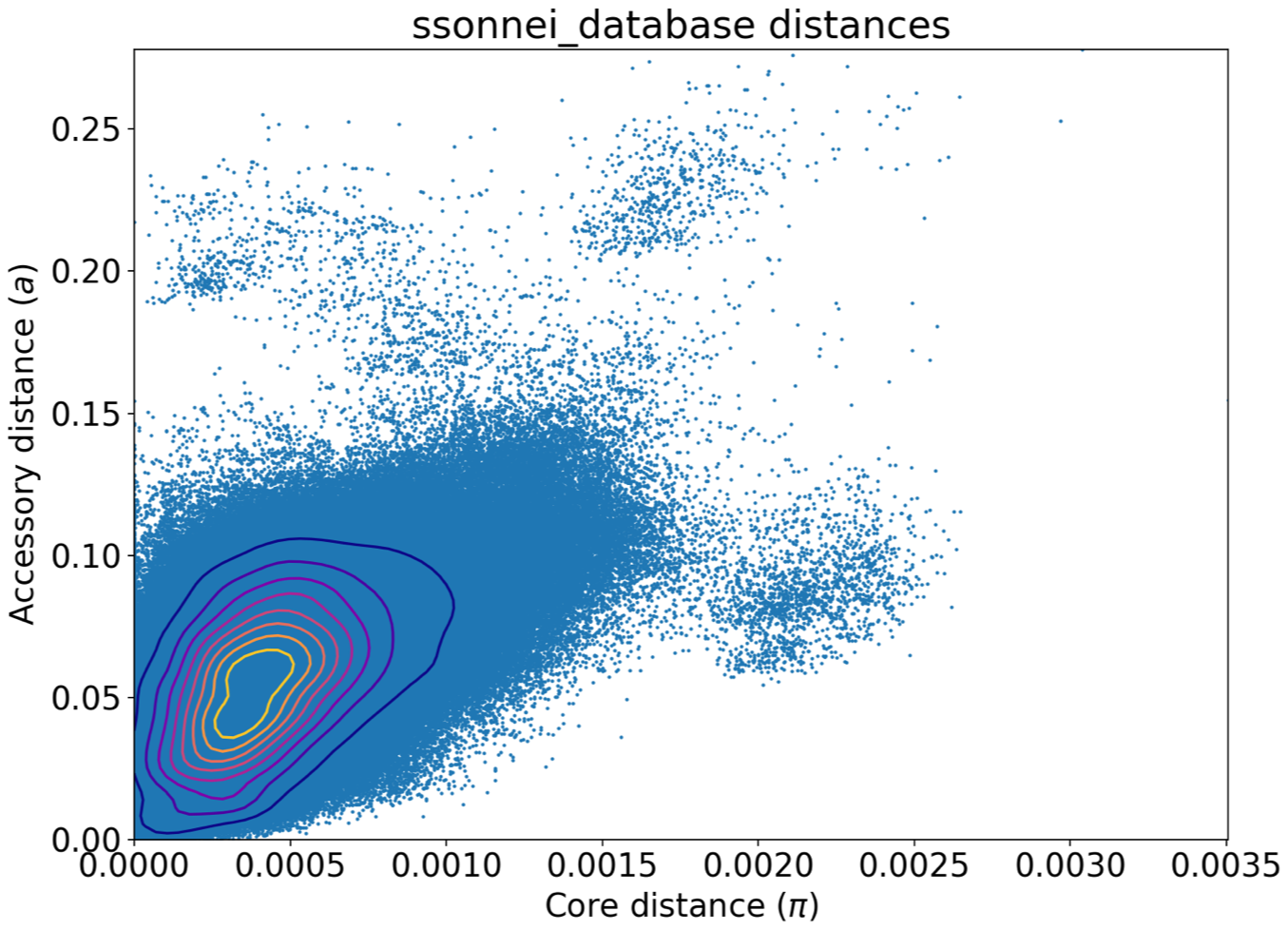

**Supplementary Figure 1**: **PopPUNK sketch of all *Shigella sonnei* isolates from UKHSA routine genomic surveillance.** PopPUNK (Lees et al., 2019) pairwise distances for all *S. sonnei* samples presented as a scatter of accessory distance (a) versus core distance (π). Points are overlaid with 2-D kernel-density contours highlighting the dense low-distance kernel.

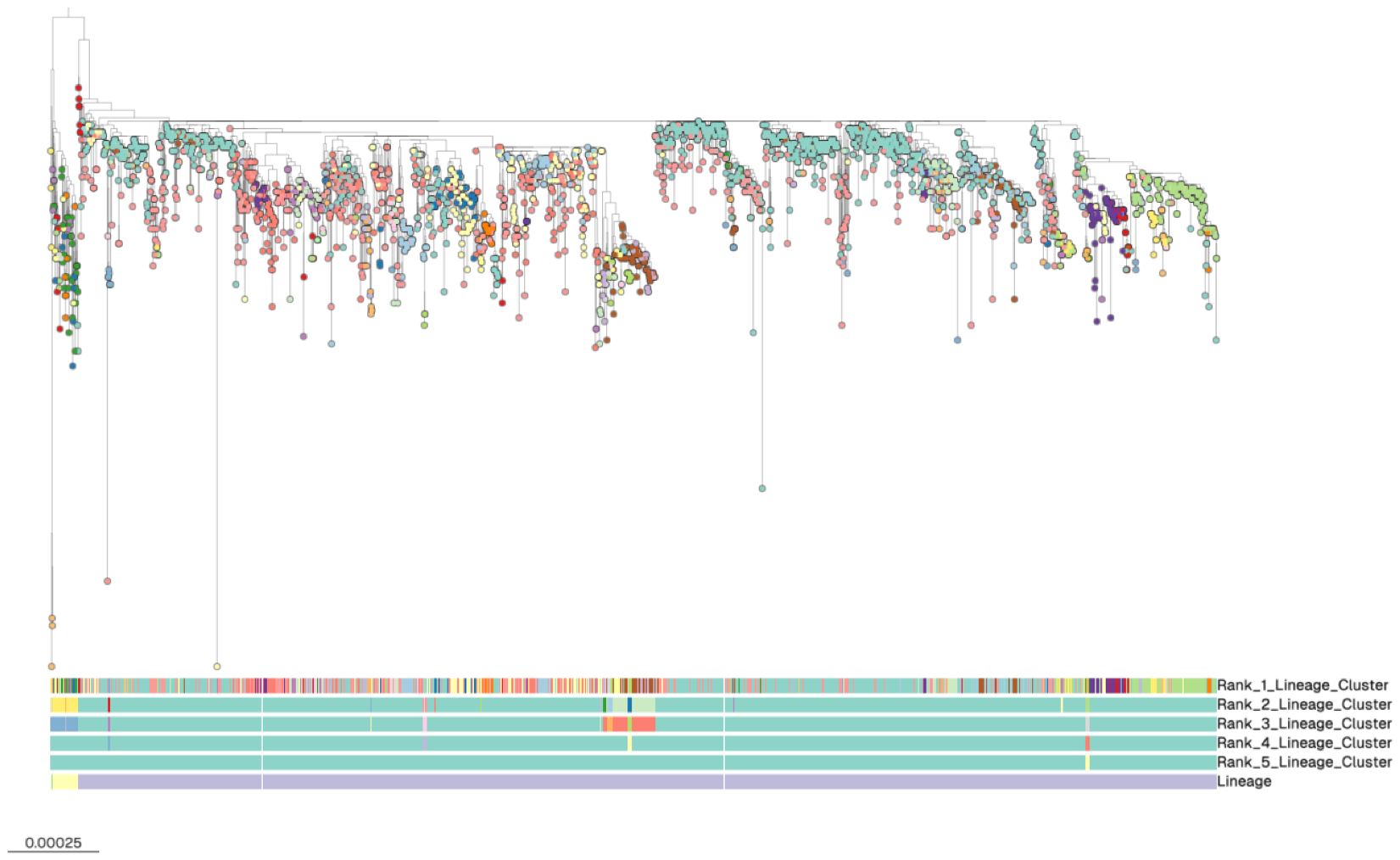

**Supplementary Figure 2**: **Hierarchical lineage structure of all *S. sonnei* resolved using iterative‑PopPUNK.** Iterative‑PopPUNK (Zhao et al., 2023) was applied with lineage‑based model fitting to infer hierarchical population structure across multiple ranks. At the finest resolution (Rank 1), 128 lineages were inferred. Progressive clustering at higher ranks reduced the number of lineages (18 at Rank 2, 11 at Rank 3, 5 at Rank 4, 2 at Rank 5, and a single lineage at Rank 6), reflecting increasingly coarse population structure. The Microreact (Argimón et al., 2016) visualisation shown was coloured by Rank_1 lineage clusters to illustrate the maximum resolution of PopPUNK‑defined genetic structure. The three lineages correspond to the global lineage 1-3 of *S. sonnei* (Hawkey et al., 2021).

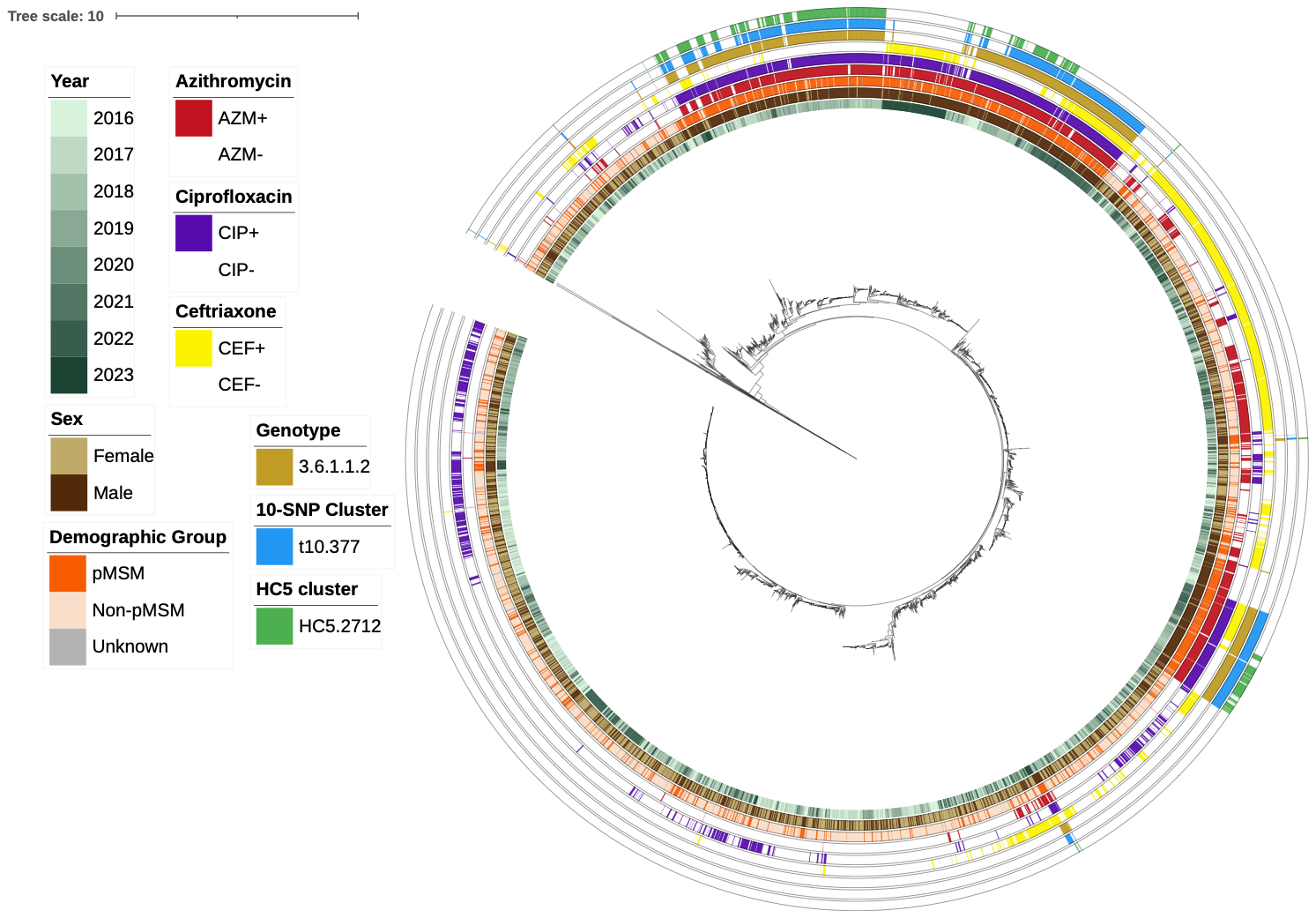

**Supplementary Figure 3**: **KPop’s neighbour-joining phylogenetic tree of 3,534 *S. sonnei* isolates from UKHSA routine genomic surveillance.** The tree was produced using KPop (Didelot and Ribeca, 2025). Metadata rings: (1) Year of isolation (2016-2023); (2) Sex (Female, Male, Unknown); (3) Demographic group (pMSM, non-pMSM, Unknown); (4) Predicted phenotypic non-susceptibility to azithromycin (AZM), ciprofloxacin (CIP), and ceftriaxone (CRO); (5) Cluster overlays for cross-scheme comparison: (i) *In silico* genotype (sonneityping): example genotype 3.6.1.1.2 highlighted (Hawkey et al., 2021); (ii) SNP-address (SnapperDB): example t10 cluster.377 highlighted (Dallman et al., 2018); (iii) cgMLST/HierCC: example HC5 cluster.2712 highlighted (Achtman et al., 2022; Zhou et al., 2021). Missing data are rendered as blank segments.

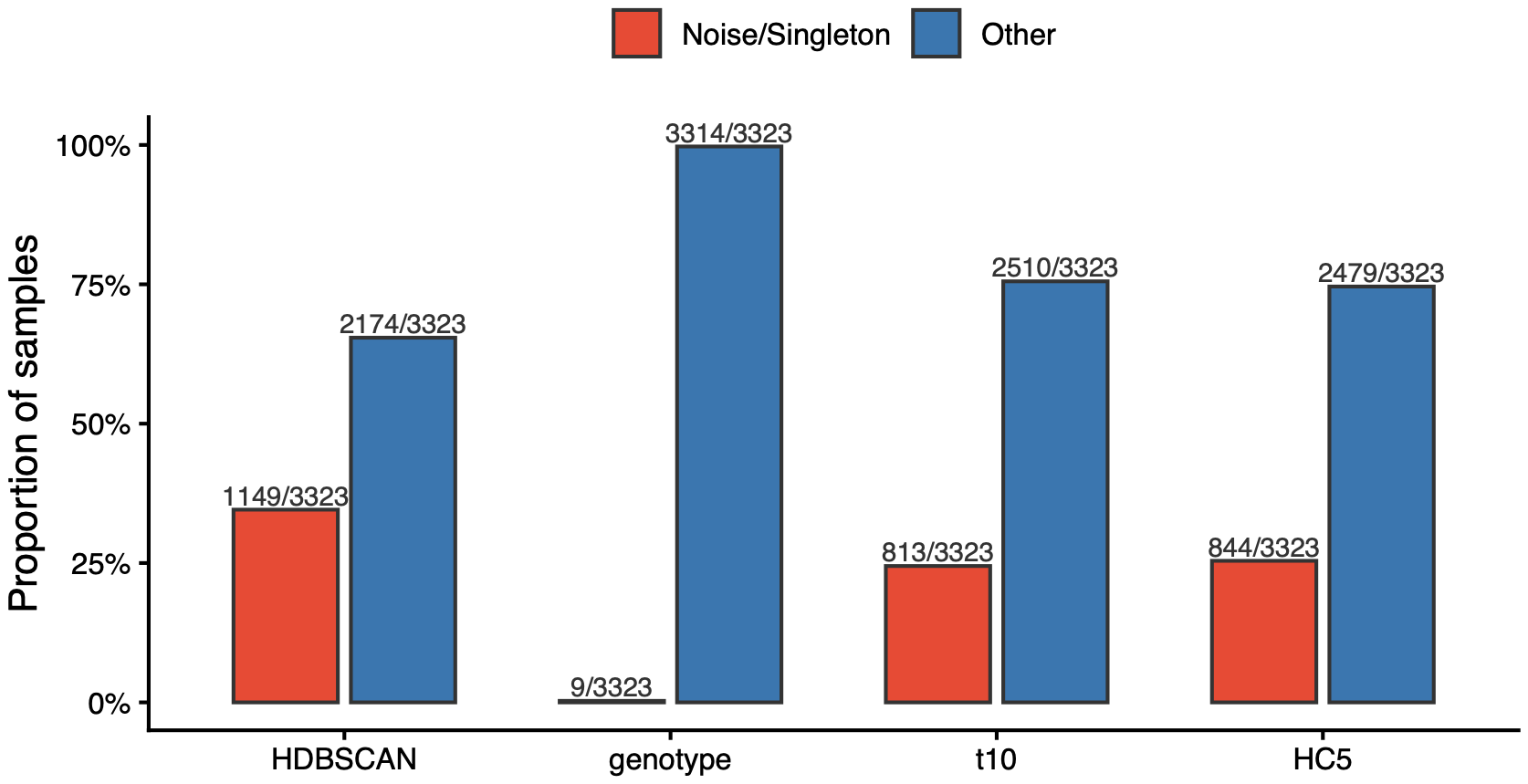

**Supplementary Figure 4**: **Proportion of *S. sonnei* samples classified as either noise or singleton by each subtyping scheme.** Bar chart showing proportion of samples classified as noise by HDBSCAN performed over KPop distances, in context with the proportion of singleton samples (only one sample assigned into the cluster) in other subtyping schema.

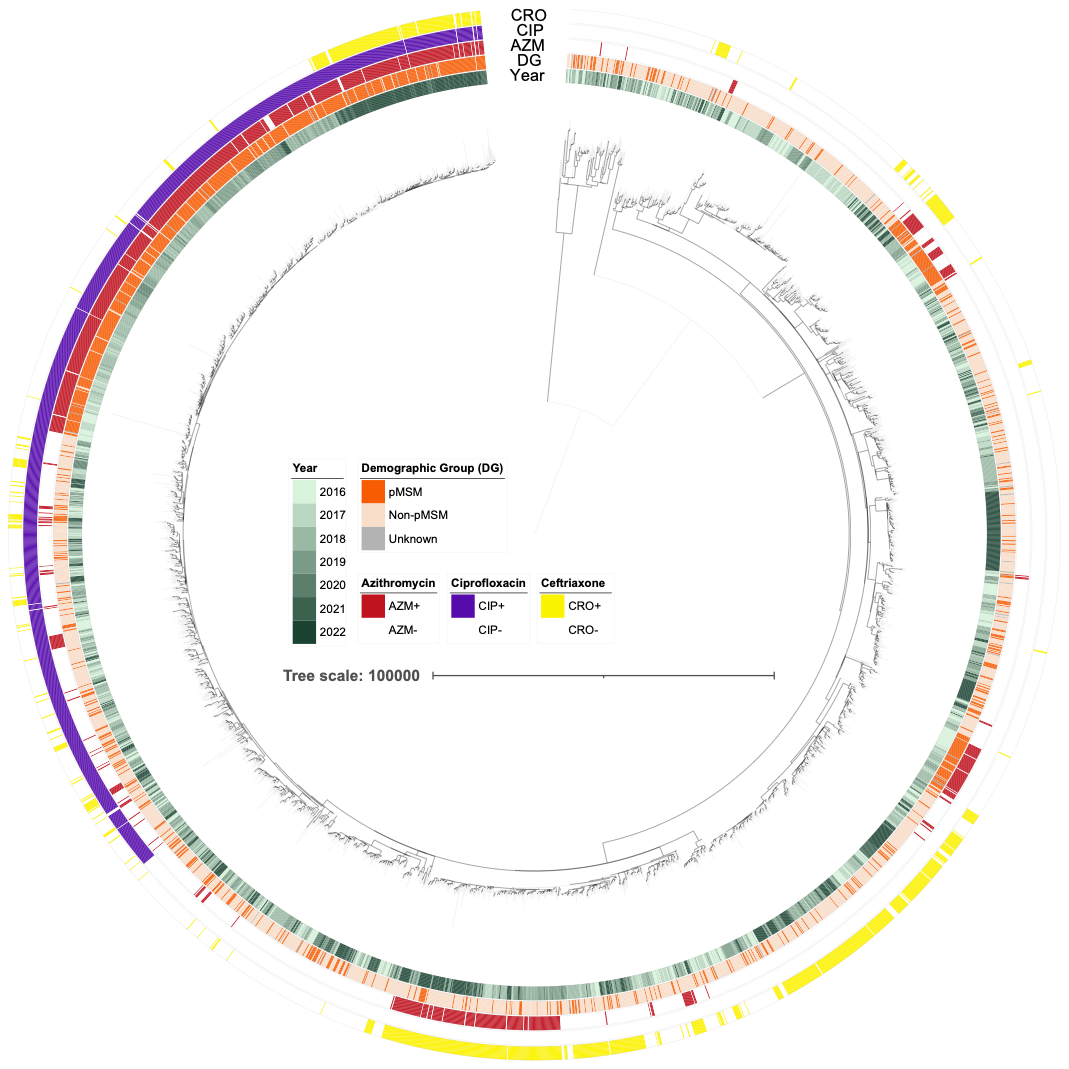

**Supplementary Figure 5**: **Maximum-likelihood phylogenetic tree of 3,534 *S. sonnei* isolates from UKHSA routine genomic surveillance.** The tree was inferred from a core-SNP alignment (S53G reference) using IQ-TREE (Nguyen et al., 2015) with the best-fit model (MFP + GTR + ASC) and 1,000 bootstrap replicates. Recombination was detected and masked with Gubbins (Croucher et al., 2015) before final tree reconstruction. Branches with support ≥ 70% are bold. Metadata rings: (1) Year of isolation (2016-2022); (2) Demographic group (pMSM, non-pMSM, Unknown); (3) Predicted phenotypic non-susceptibility to azithromycin (AZM), ciprofloxacin (CIP), and ceftriaxone (CRO). Missing data are rendered as blank segments.

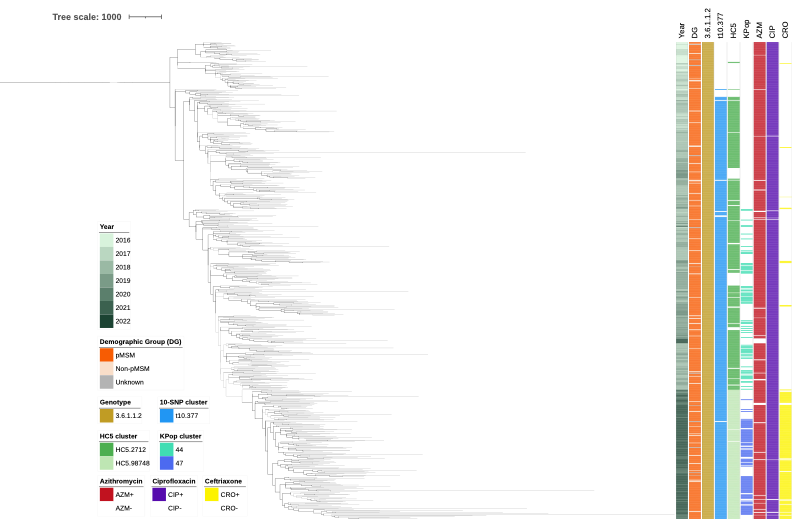

**Supplementary Figure 6**: **Truncated Genotype 3.6.1.1.2 isolates (n = 683) from maximum-likelihood phylogenetic tree of 3,534 S. sonnei isolates.** The tree was inferred from a core-SNP alignment (S53G reference) using IQ-TREE (Nguyen et al., 2015) with the best-fit model (MFP + GTR + ASC) and 1,000 bootstrap replicates. Recombination was detected and masked with Gubbins (Croucher et al., 2015) before final tree reconstruction. Branches with support ≥ 70% are bold. Metadata rings: (1) Year of isolation (2016-2022); (2) Demographic group (pMSM, non-pMSM, Unknown); (3) Cluster overlays for cross-scheme comparison: (i) *In silico* genotype (sonneityping): example genotype 3.6.1.1.2 highlighted (Hawkey et al., 2021); (ii) SNP-address (SnapperDB): example t10 cluster.377 highlighted (Dallman et al., 2018); (iii) cgMLST/HierCC: example HC5 cluster.2712 and cluster.98748 highlighted (Achtman et al., 2022; Zhou et al., 2021); (iv) KPop: example cluster 44 and 47 highlighted (Didelot and Ribeca, 2025); (4) Predicted phenotypic non-susceptibility to azithromycin (AZM), ciprofloxacin (CIP), and ceftriaxone (CRO). Missing data are rendered as blank segments.

**References**

Achtman, M., Zhou, Z., Charlesworth, J., Baxter, L., 2022. EnteroBase: hierarchical clustering of 100 000s of bacterial genomes into species/subspecies and populations. Philos. Trans. R. Soc. B Biol. Sci. 377, 20210240. https://doi.org/10.1098/rstb.2021.0240

Argimón, S., Abudahab, K., Goater, R.J.E., Fedosejev, A., Bhai, J., Glasner, C., Feil, E.J., Holden, M.T.G., Yeats, C.A., Grundmann, H., Spratt, B.G., Aanensen, D.M., 2016. Microreact: visualizing and sharing data for genomic epidemiology and phylogeography. Microb. Genomics 2, e000093. https://doi.org/10.1099/mgen.0.000093
